# Effect of starting metformin on severe and long COVID-19 among people living with type 2 diabetes in England: a new user cohort study in OpenSAFELY-TPP

**DOI:** 10.64898/2026.11.09.26367638

**Authors:** Alain Amstutz, Venexia Walker, Paul Madley-Dowd, Rachel Denholm, Mengxuan Zou, Tom Palmer, OpenSAFELY Collaborative, Sophie V Eastwood, William J Hulme, Jonathan A C Sterne

**Author notes:** **E-mail addresses:** VW.

## Abstract

**Introduction:** Observational studies report lower COVID-19 mortality among metformin users, but most used prevalent-user designs comparing long-term survivors with heterogeneous controls. Randomised trials suggest little or no mortality effect. No large observational study has used a new-user design to estimate the effect of starting versus not starting metformin on severe and Long COVID outcomes in adults with type 2 diabetes (T2D). We aimed to estimate this effect on COVID-19-related hospitalisation or death, COVID-19 diagnoses, and Long COVID among adults with T2D in England.

**Research design and methods:** Using OpenSAFELY-TPP linked NHS electronic health records, we conducted a population-based cohort study with a landmark new-user design. Adults aged 18-85 years newly diagnosed with T2D (August 2018-August 2019) were classified as starting metformin monotherapy within 6 months or not starting any antidiabetic drug, based on prescription data. Follow-up began 6 months after diagnosis and before the UK pandemic (avoiding immortal time) for up to 2 years. We estimated intention-to-treat and per-protocol effects using weighted Cox and pooled logistic regression, with negative (bone fracture) and positive (diabetes-related death) control outcomes.

**Results:** Of 52,341 eligible people, 19,599 (37.4%) started metformin. For COVID-19-related hospitalisation or death, the maximally adjusted hazard ratio (HR) of metformin versus no antidiabetic was 1.17 (95% CI 1.02-1.35; 2-year risk difference 0.36%, 95% CI 0.09-0.62; intention-to-treat). The per-protocol HR for COVID-19-related hospitalisation or death was 1.11 (95% CI 0.96-1.29). The Long COVID estimate was imprecise (HR 1.39, 95% CI 0.83-2.33; intention-to-treat). For bone fracture (negative control) the estimate was null (HR 1.00), but for diabetes-related deaths (positive control) it was elevated (HR 1.41, 95% CI 1.09-1.81), indicating unmeasured confounding.

**Conclusions:** The risks of severe and long COVID were slightly higher among people who started metformin, compared with those who did not start. These results plausibly reflect unmeasured confounding. These findings align more closely with randomised evidence than earlier prevalent-user studies and argue against a clinically meaningful benefit of starting metformin for COVID-19 prevention in newly diagnosed T2D.

**What is already known on this topic:** Observational studies and their meta-analyses have consistently reported 30-40% lower COVID-19 mortality among metformin users, yet most used prevalent-user designs that compare long-term survivors with mixed control groups and can overstate benefit. Randomised trials suggest little or no effect on mortality.

**What this study adds:** Using a new-user design in more than 52,000 adults with newly diagnosed T2D, risks of severe and long COVID were slightly higher among people who started metformin, compared with those who did not start. These results plausibly reflect unmeasured confounding rather than harm.

**How this study might affect research, practice or policy:** These findings align with randomised evidence and argue against a clinically meaningful benefit of starting metformin for preventing severe or Long COVID in people with T2D. They also illustrate how new-user designs can reconcile discrepant observational and trial results and should be prioritised in future drug-repurposing studies.

**Lay summary:** Metformin is a medication commonly used to treat type 2 diabetes. During the COVID-19 pandemic, some studies suggested it might also protect against severe COVID-19, but many such studies had design limitations that could have made metformin look more beneficial than it truly is. In this study, we used NHS health records from over 52,000 adults in England who were newly diagnosed with type 2 diabetes to examine whether starting metformin shortly after diagnosis reduced the risk of being hospitalised or dying from COVID-19, being diagnosed with COVID-19. We found no evidence that starting metformin protected against any of these outcomes, although the effect on long COVID was imprecisely estimated. Our findings are consistent with results from clinical trials and suggest that metformin should not be expected to prevent severe COVID-19 in people with type 2 diabetes.

## Introduction

During the COVID-19 pandemic several drug repurposing candidates, including metformin, were investigated for their effect on COVID-19 outcomes.^1^ Metformin is the most commonly prescribed first-line treatment for type 2 diabetes (T2D) and is an inexpensive, readily accessible medication with a well-known safety profile. It has been hypothesized that metformin helps in preventing severe and long COVID-19 outcomes due to its anti-inflammatory and potential antiviral properties.^2,3^

Several randomised trials, summarized in a recent meta-analysis, have evaluated the use of metformin to prevent severe COVID-19 outcomes among a broad at-risk population.^4^ The meta-analysis concluded that metformin has little to no effect on mortality, but some (uncertain) effect on severe COVID-19 outcomes. This is in contrast to results from observational studies and meta-analyses thereof, which generally reported 30-40% lower odds of mortality among metformin users compared with non-users.^5–8^ Importantly, most observational studies used prevalent-user designs to assess the effect of metformin rather than studying the effect of starting, compared with not-starting, metformin. Prevalent users are individuals who remain on long-term metformin therapy so are, by definition, survivors who tolerated the drug and maintained engagement with healthcare. Therefore, comparing prevalent-users with non-users can over-estimate the benefit of treatment as former-users, who are less likely to have benefitted, are excluded. For Long COVID, the evidence is more consistent: both randomised and observational analyses reported a protective effect of metformin on incident Long COVID.^9^

To our knowledge, no large observational study focusing on adults with T2D has used a new-user design to estimate the effect of starting, compared with not starting, metformin among people with T2D on subsequent severe and Long COVID-19 outcomes. Such a design more closely mirrors the causal question addressed in randomised trials. If metformin does confer protection, it would represent a widely available, low-cost intervention in a population at elevated risk of severe COVID-19.

We aimed to estimate the effect of starting versus not starting metformin on COVID-19 related hospitalisations or death, COVID-19 diagnoses, and Long COVID diagnoses, among adults living with T2D in England.

## Research Design and Methods

### Data sources

All data were linked, stored and analysed securely using the OpenSAFELY platform^10^, https://www.opensafely.org/, as part of the NHS England OpenSAFELY Analytics Service (https://github.com/OpenSAFELY-Core). Primary care records managed by the GP software provider, TPP, were linked to the Office for National Statistics (ONS) death registration data, the national coronavirus testing records from the Second Generation Surveillance System (SGSS), the national vaccine register (National Immunisation Management System), and the NHS Secondary Use Service (SUS) data, through OpenSAFELY. OpenSAFELY-TPP contains pseudonymised data of approximately 40% of the English population, including coded diagnoses, medications and physiological parameters. No data from patients who registered a Type-1 Opt Out with their GP surgery were included in this study. More information on the patients available for selection in the Service is available in the OpenSAFELY data page of the documentation. The analysis dataset was defined and created using ehrQL (v1.0) / Python (v3.10). Data management and analyses were conducted in R version 4.4.3. For disclosure control, all counts were midpoint-6 rounded (https://docs.opensafely.org/outputs/sdc/). The protocol, codelists and analysis code are shared openly for review and re-use under MIT open license (https://github.com/opensafely/metformin_covid/)

### Study design and participants

We used a population-based cohort design (Figure 1). People with a T2D diagnosis between August 1st, 2018, and August 1st, 2019 (‘diagnosis window’) were classified as starting or not starting metformin within 6 months after T2D diagnosis. Follow-up started at 6 months after diagnosis (‘landmark approach’). Because the latest possible date of T2D diagnosis (August 1st, 2019) was 6 months before the start of the pandemic in the UK (February 1st, 2020), no COVID-related outcomes could occur between T2D diagnosis and the start of follow-up. Therefore, our design avoided creating ‘immortal time’ during which outcomes were excluded from analyses.^11^ Follow-up ended at the earliest of: outcome, death, end of registration, maximum follow-up period (2 years) or administrative end of study (March, 1st, 2022 [end of systematic SARS-CoV-2 mass testing in the UK]). People who died or were lost to follow-up (deregistration from the general practice) within 6 months after T2D diagnosis were excluded from analyses.

**Figure 1.**
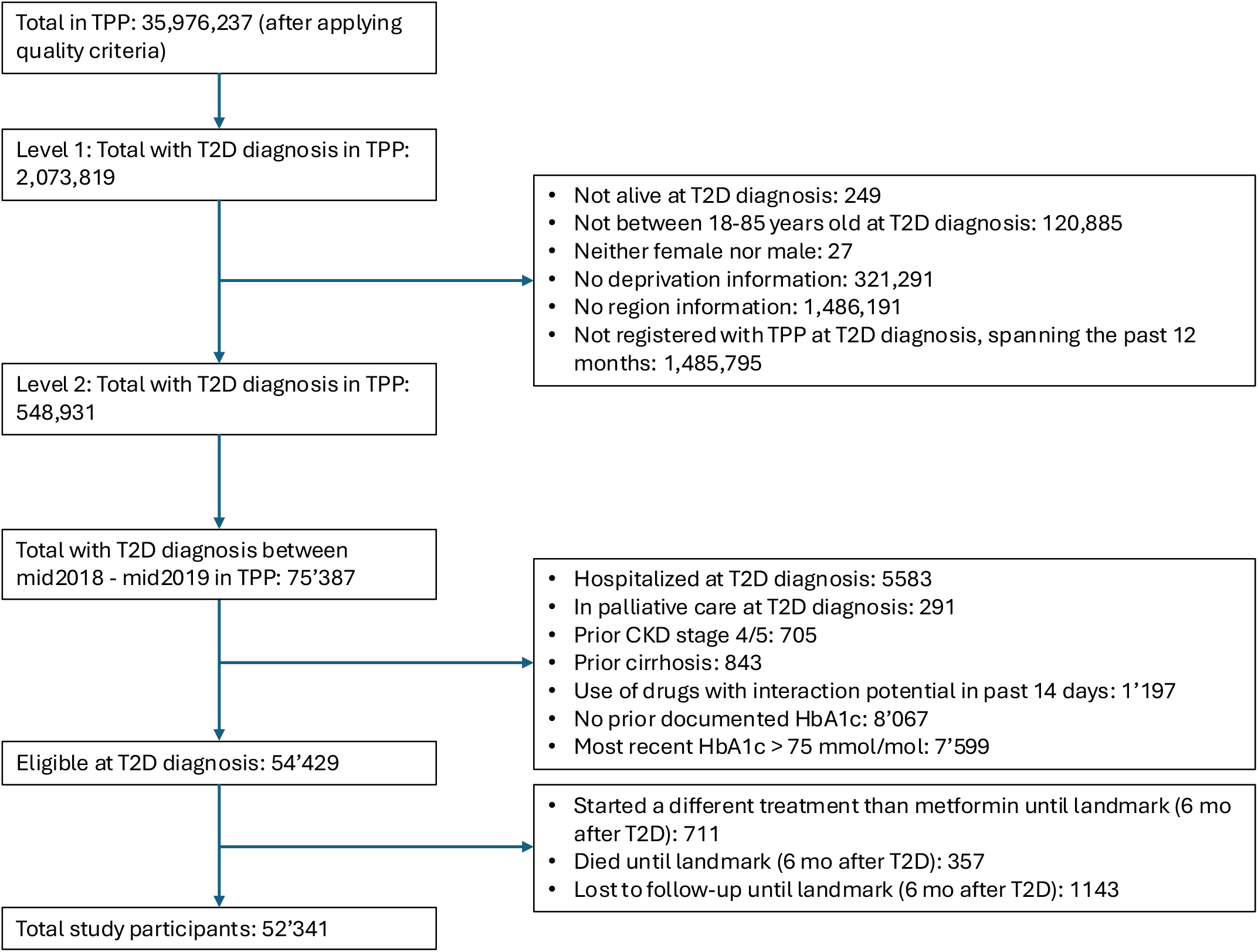
Illustration of the study design

Eligible people had a T2D diagnosis within the diagnosis window, had been registered for at least 12 months before diagnosis with an English GP using TPP software, were aged between 18 years and 85 years at the time of T2D diagnosis, and had available data for sex, region, and area deprivation. People hospitalized, in palliative care or a nursing home, with a missing glycated hemoglobin (HbA1c) or a HbA1c above 75 mmol/mol in the 2 years prior to T2D diagnosis were excluded. We excluded people with known history of hypersensitivity to metformin, moderate to severe renal impairment, or advanced decompensated liver cirrhosis, as well as people with prior use of metformin or another glucose-lowering medication (SGLT2, DPP4, sulfonylurea, thiazolidinedione, GLP-1, meglitinides, alpha-glucosidase-inhibitors, insulin) or any medication with metformin drug-drug-interaction potential (Cimetidine, hydroxychloroquine, tafenoquine, patiromer, ranolazine, Monoamine Oxide Inhibitors, Alpha-1 antagonists, Sotalol, Clonidine, Phosphodiesterase 5 inhibitors, Methyldopa, Prazosin, Terasozin, Doxazosin) in the past 14 days prior to T2D diagnosis (appendix text S1).

This study is reported according to the REporting of studies Conducted using Observational Routinely collected health Data for PharmacoEpidemiological research (RECORD-PE) reporting guideline^12^ (checklist in appendix text S2) and was approved by the Health Research Authority (REC reference 22/PR/0095) and by the University of Bristol’s Faculty of Health Sciences Ethics Committee (reference 117269). The study protocol, including amendments and track changes, is available in the GitHub repository (https://github.com/opensafely/metformin_covid/tree/main/protocol).

### Treatment strategies

We compared the treatment strategies ‘start metformin monotherapy (i.e. no other antidiabetic drug) within 6 months of T2D diagnosis and continue metformin during follow-up unless a contraindication occurs’ and ‘do not start metformin or any other antidiabetic drug within 6 months of T2D diagnosis or any time during the follow-up’. The exact drugs of interest are outlined in appendix table S1.

Participants who started any antidiabetic drug other than metformin within 6 months of T2D diagnosis were excluded from analyses

### Outcomes

The primary outcome was a composite of COVID-19-related hospitalisation and COVID-19-related death. COVID-19-related hospitalisations, obtained from secondary care data including emergency care attendance, were defined as any (emergency) hospitalisation listing a COVID-19 diagnosis in any position. Deaths were identified using linked ONS death registry data. COVID-19-related death was defined as a death where the underlying or contributory cause on the death certificate was COVID-19. If a person was hospitalised before death, the date of hospitalisation was used. The secondary outcome ‘COVID-19 diagnosis’ was defined using an established algorithm combining any event indicating a COVID-19 diagnosis (i.e., a positive SARS-CoV-2 test, a COVID-19 diagnostic code in primary or secondary care, or a COVID-19-related death) in SGSS, SUS and ONS death registry.^13^ The secondary outcome ‘Long COVID’ was defined using Long COVID diagnostic codes in primary care, introduced by the National Health Services in the UK, supplemented by viral fatigue codes.^14^ In sensitivity analyses, we used diabetes-related deaths as a positive control outcome and hospitalisations due to bone fracture as a negative control outcome (appendix table S2).

### Covariates and confounders

Covariates identified as potential confounders included age (splines with knot locations based on 10, 50 and 90 percentile), baseline sex (binary), ethnicity (categorised as White; Black or Black British; Asian or Asian British; Other, including Missing and Mixed), deprivation (using the Index of Multiple Deprivation [IMD] - the official measure of relative deprivation for small areas in England defined by 7 domains, including income and barriers to housing and services; categories as 1 [most deprived] to 5 [least deprived]), rural/urban area (binary), smoking status (categorised as Never; Ever, including Missing; Current), health care worker (binary), GP-patient interactions in the year prior to T2D diagnosis (continuous), calendar time of T2D diagnosis (continuous), number of HbA1c measurements in previous 2 years (continuous) - all assessed at baseline. Additional baseline covariates, which were also time-updated over the follow-up, included HbA1c (continuous), total cholesterol/high-density lipoprotein cholesterol ratio (categorised as below 3.5:1; 3.5:1 to 5:1; above 5:1; missing), prediabetes, diabetes-related complications, and binary indicators for fifteen comorbidities (appendix text S3 and appendix figure S1). Region was treated as a stratification factor (categorised into 7 regions: East, London, Midlands, North East and Yorkshire, North West, South East, South West).

### Causal estimands and statistical analyses

We described demographic and clinical characteristics at T2D diagnosis date. We generated a propensity score for receiving metformin for each individual, defined by the baseline covariates listed above, and examined how this differed by receipt of metformin.

We estimated both ‘intention-to-treat’ (ITT) and ‘per-protocol’ effects comparing the two treatment strategies. To estimate the observational analogue of the ITT effect, we compared people who started metformin monotherapy within 6 months of T2D diagnosis with people who did not start metformin monotherapy or any other antidiabetic drug within 6 months of T2D diagnosis. We fitted Cox models with calendar time scale and start of follow up 6 months after T2D diagnosis (the landmark date).

Follow-up ended at the earliest of the outcome, death or practice deregistration. For each outcome, we estimated age- and sex-adjusted and maximally adjusted hazard ratios comparing the treatment strategies. All models were stratified by region. We estimated 2-year hazard ratios (HRs) with standard Wald 95% confidence intervals. In addition, we used an inverse probability of treatment-weighted Kaplan-Meier approach to estimate the marginal (population-average) cumulative incidence of the primary outcome for each treatment group. From these, we estimated 2-year risk differences (RDs) and risk ratios (RRs) with 95% confidence intervals using the delta method.

To estimate the per-protocol effect, we artificially censored people in the no antidiabetic group who deviated from their assigned treatment strategy by starting metformin or any other antidiabetic more than 6 months after diagnosis of T2D. We used inverse probability of treatment (with metformin or other antidiabetic drugs) weighting to account for this informative censoring. To operationalize this, we discretised person-time into monthly intervals containing information on the time-updated covariate values at the beginning of the interval, treatment status during the interval, and whether the outcome event occurred in the next interval. We used pooled logistic regression to estimate each person’s probability of being treated in each interval, based on the baseline and time-updated covariates. These were used to derive each person’s cumulative probability of remaining uncensored during each time interval. We calculated stabilised weights, for which the numerator was the probability of receiving the treatment that each individual received given their past treatment history (treatment, time variables, and baseline HbA1c as the strongest predictor), and we truncated them at the 1st and 99th percentiles (sensitivity analysis without truncation were also conducted). We also used inverse probability of censoring weights, based on the same time-varying covariates, to account for potentially informative censoring due to deregistration from the general practice.

Finally, we fitted the outcome model, using pooled logistic regression on the discretised dataset, weighted by the product of the two weights above, and including linear and quadratic terms for month of follow-up to model the baseline hazard. We fitted the outcome model with an interaction between time (month and month^2^) and treatment. We obtained 2-year HRs, cumulative incidence curves, 2-year RDs and RRs, with 95% confidence intervals using bootstrapping.

We conducted subgroup analyses on the primary outcome by age group (below vs ≥60 years), sex (female vs male), ethnicity (white vs all other ethnicities combined), IMD (most deprived quintile vs other), BMI (obese vs overweight vs low/normal weight or missing), and HbA1c (below vs ≥59 mmol/mol). We used Cochran’s Q statistic to assess the differences within subgroups.

We conducted a sensitivity analysis restricting the study population to the propensity score region with substantial overlap (0.1-0.8).

## Results

We identified 54,341 people with T2D who met the inclusion criteria (figure 2). Between T2D diagnosis and landmark (6 months later), 357 (0.7%) died, 1,143 (2.1%) were lost to follow-up and 711 (1.3%) started an antidiabetic treatment other than metformin. Therefore, 52,341 people were included in the analyses. Among these, 19,599 (37.4%) were prescribed metformin within 6 months of T2D diagnosis. People who were prescribed metformin were younger (median age 58 [49-68] versus 64 [54-73] years) and had fewer comorbidities (for example, arterial hypertension: 9,441 [48%] versus 18,027 [55%]) than the no antidiabetic group (table 1). However, they had higher TC/HDL ratio (5:1 ratio category and above: 4,131 [21%] versus 4,953 [15%]) and higher HbA1c (59-75 mmol/mol category: 5,595 [29%] versus 1,659 [5%]). Other characteristics were broadly similar across the groups.

**Table 1.** Characteristics at T2D diagnosis.

| **Characteristic** | **Metformin** | **No antidiabetic** |
|---|---|---|
|  | **19,599** | **32,739** |
| **Age**, median (IQR), mean (SD) | 58 (49, 68); 58 (13) | 64 (54, 73); 63 (13) |
| **Age groups** |  |  |
| 18-39 | 1,599 (8.2%) | 1,281 (3.9%) |
| 40-59 | 9,195 (46.9%) | 11,013 (33.6%) |
| 60-79 | 7,941 (40.5%) | 17,175 (52.5%) |
| 80 or older | 861 (4.4%) | 3,267 (10%) |
| **Sex** |  |  |
| Female | 8,949 (45.7%) | 14,949 (45.7%) |
| Male | 10,647 (54.3%) | 17,787 (54.3%) |
| **Ethnicity** |  |  |
| White | 15,261 (77.9%) | 26,655 (81.4%) |
| Asian | 2,889 (14.7%) | 3,975 (12.1%) |
| Black | 639 (3.3%) | 915 (2.8%) |
| Other | 807 (4.1%) | 1,197 (3.7%) |
| **Deprivation** |  |  |
| 1 (most deprived) | 5,613 (28.6%) | 7,197 (22%) |
| 2 | 4,647 (23.7%) | 6,987 (21.3%) |
| 3 | 3,975 (20.3%) | 7,215 (22%) |
| 4 | 3,051 (15.6%) | 6,111 (18.7%) |
| 5 (least deprived) | 2,313 (11.8%) | 5,229 (16%) |
| **Region** |  |  |
| East | 4,599 (23.5%) | 6,465 (19.7%) |
| London | 1,983 (10.1%) | 2,391 (7.3%) |
| Midlands | 4,041 (20.6%) | 6,687 (20.4%) |
| North East and Yorkshire | 4,119 (21%) | 6,801 (20.8%) |
| North West | 1,857 (9.5%) | 3,573 (10.9%) |
| South East | 1,185 (6%) | 2,295 (7%) |
| South West | 1,815 (9.3%) | 4,527 (13.8%) |
| **Rural/urban** |  |  |
| Urban conurbation | 6,681 (34.1%) | 9,945 (30.4%) |
| Urban city or town | 9,723 (49.6%) | 15,849 (48.4%) |
| Rural town or village | 3,195 (16.3%) | 6,945 (21.2%) |
| **Smoking status** |  |  |
| Smoker | 3,723 (19%) | 4,761 (14.5%) |
| Ever | 8,349 (42.6%) | 15,093 (46.1%) |
| Never | 7,527 (38.4%) | 12,885 (39.4%) |
| **Healthcare worker** | 489 (2.5%) | 669 (2%) |
| **Consultation rate in previous year, median (IQR), mean (SD)** | 8 (5, 13); 11 (9) | 9 (5, 14); 11 (9) |
| **Body Mass Index (BMI) categories** |  |  |
| Underweight, healthy, or unknown | 6,309 (32.2%) | 10,845 (33.1%) |
| Overweight (25-29.9) | 3,147 (16.1%) | 7,011 (21.4%) |
| Obese (>30) | 10,137 (51.7%) | 14,883 (45.5%) |
| **HbA1c in mmol/mol**, median (IQR), mean (SD) | 53 (50, 60); 55 (8) | 49 (48, 52); 50 (5) |
| **HbA1c categories in mmol/mol** |  |  |
| below 42 | 237 (1.2%) | 1,341 (4.1%) |
| 42-58 | 13,767 (70.2%) | 29,745 (90.9%) |
| 59-75 | 5,595 (28.5%) | 1,659 (5.1%) |
| **TC/Chol ratio,** median (IQR), mean (SD) | 4.24 (3.44, 5.17); 4.41 (1.39) | 3.90 (3.15, 4.78); 4.06 (1.26) |
| **TC/HDL ratio categories** |  |  |
| below 3.5:1 | 4,197 (21.4%) | 9,759 (29.8%) |
| 3.5:1 to 5:1 | 7,413 (37.8%) | 11,985 (36.6%) |
| above 5:1 | 4,131 (21.1%) | 4,953 (15.1%) |
| Unknown | 3,855 (19.7%) | 6,039 (18.4%) |
| **History of acute myocardial infarct** | 1,407 (7.2%) | 2,835 (8.7%) |
| **History of stroke** | 651 (3.3%) | 1,329 (4.1%) |
| **History of other arterial embolism** | 45 (0.2%) | 123 (0.4%) |
| **History of venous thromboembolism** | 615 (3.1%) | 1,125 (3.4%) |
| **History of heart failure** | 837 (4.3%) | 2,025 (6.2%) |
| **History of angina pectoris** | 1,875 (9.6%) | 4,191 (12.8%) |
| **History of dementia** | 183 (0.9%) | 459 (1.4%) |
| **History of cancer** | 5,709 (29.1%) | 9,279 (28.3%) |
| **History of arterial hypertension** | 9,441 (48.2%) | 18,027 (55.1%) |
| **History of depression** | 7,521 (38.4%) | 11,007 (33.6%) |
| **History of COPD** | 1,629 (8.3%) | 3,063 (9.4%) |
| **History of liver disease** | 153 (0.8%) | 243 (0.7%) |
| **History of CKD** | 1,515 (7.7%) | 4,143 (12.7%) |
| **History of PCOS** | 27 (0.1%) | 21 (0.1%) |
| **History of prediabetes** | 12,363 (63.1%) | 23,937 (73.1%) |
| **Diabetes complication** | 99 (0.5%) | 351 (1.1%) |
| **Number of HbA1c measurements, previous 2y**, median (IQR), mean (SD) | 2.00 (2.00, 4.00); 2.93 (1.86) | 3.00 (2.00, 4.00); 3.14 (1.87) |
| **Number of lifestyle advices, previous 2y**, median (IQR), mean (SD) | 0.00 (0.00, 1.00); 0.54 (1.05) | 0.00 (0.00, 1.00); 0.64 (1.21) |

**Figure 2.**
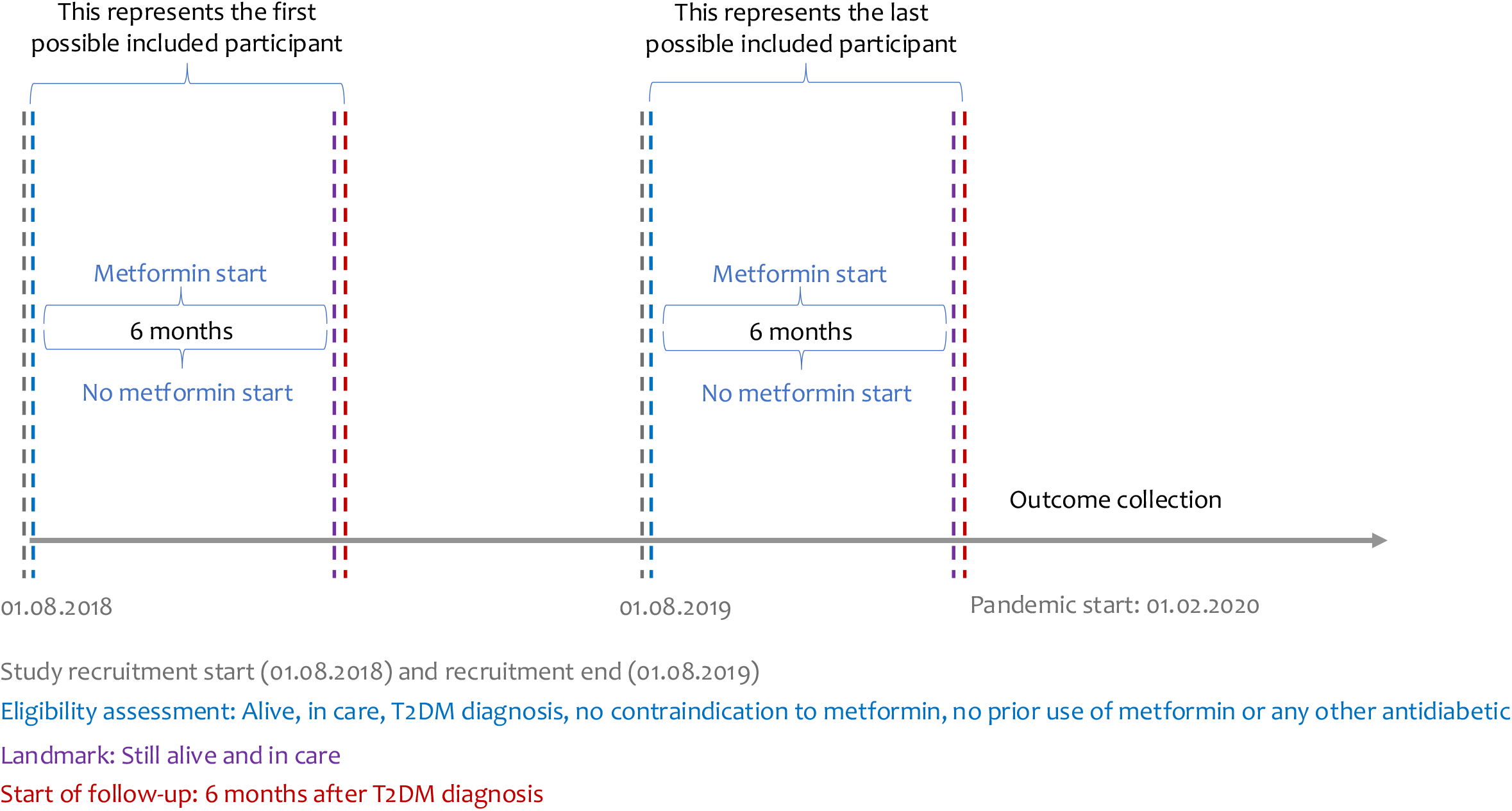
Flowchart

The densities of the propensity score for those prescribed and not prescribed metformin had peaks at 0.3 and 0.25, respectively (appendix figure S2). There was reasonable overlap between the groups across the entire propensity score distribution. Characteristics of people who dropped out due to death or loss to follow-up before the landmark and before pandemic start were similar to those of people included in the study population (appendix table S3 and appendix table S4).

### Intention-to-treat analysis of the primary outcome

Overall, 375 (1.92%) of 19,599 participants in the metformin group experienced a COVID-19-related hospitalisation or death by 2 years follow-up, compared with 597 (1.82%) of 32,739 participants in the no antidiabetic group. The cumulative incidence of COVID-19-related hospitalisation or death was 2.23% (95% CI 2.02% to 2.45%) in the metformin group and 1.87% (95% CI 1.72% to 2.03%) in the no antidiabetic group, with an absolute cumulative RD of 0.36% (95% CI 0.09% to 0.62%) and RR of 1.19 (95% CI 1.05 to 1.35) at maximum follow-up (appendix figure S4). The Incidence of COVID-19-related hospitalisation or death was higher in the metformin group (maximally adjusted HR 1.17, 95% CI 1.02 to 1.35; figure 3). Restricting the study population to the propensity score region with substantial overlap yielded an adjusted HR of 1.12 (95% CI 0.97 to 1.30; (figure 3)) on COVID-19-related hospitalisations or death.

**Figure 3.**
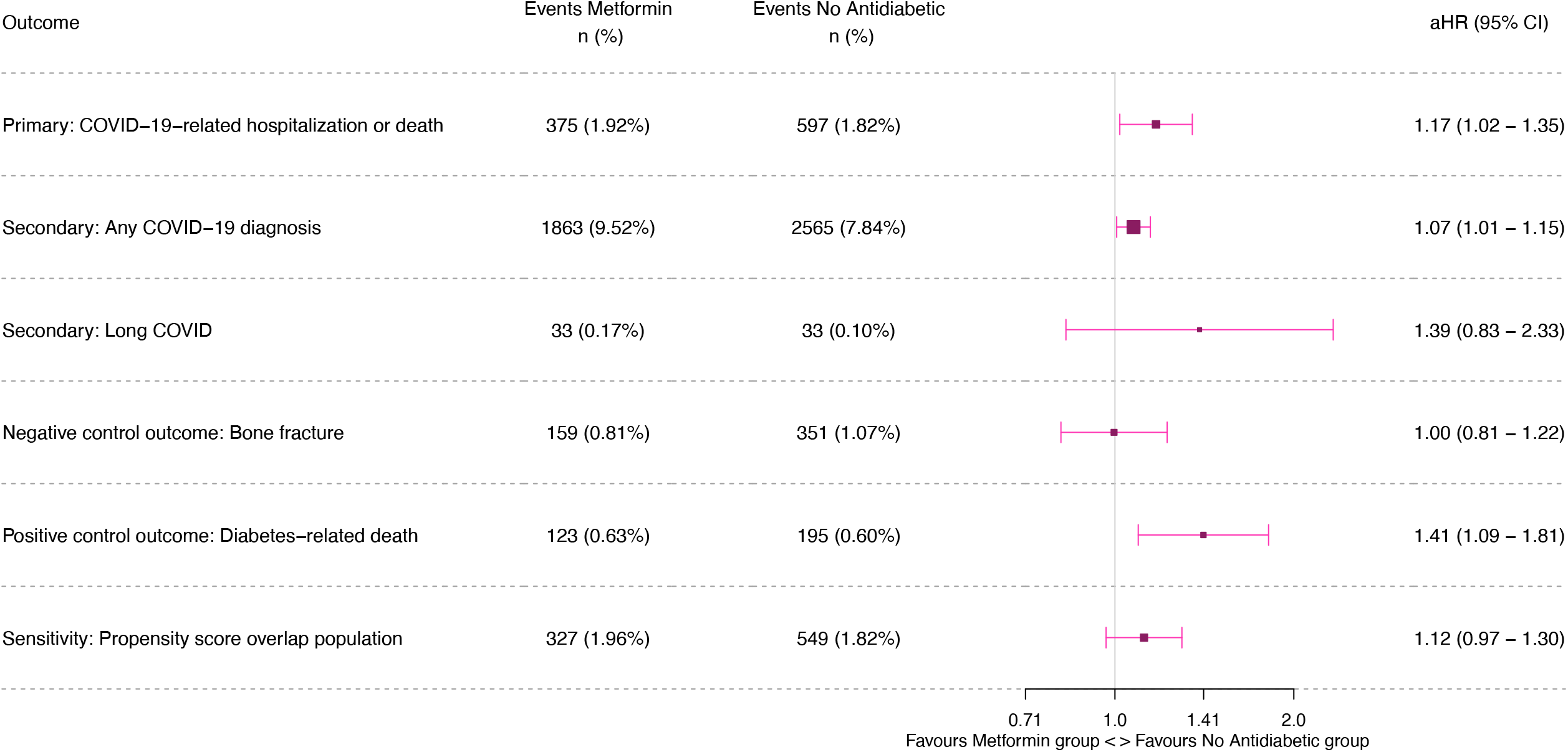
All outcomes, maximal adjusted model, primary analysis (Cox model, intention-to-treat) Footnote: Hazard ratio, adjusted for age, sex, ethnicity, deprivation, rural/urban area, smoking status, health care worker, GP-patient interactions in the year prior to T2D diagnosis, calendar time of T2D diagnosis, Number of HbA1c measurements, HbA1c, total cholesterol/high-density lipoprotein cholesterol ratio, prediabetes, diabetes-related complications, and binary indicator for fifteen comorbidities; abbreviations: aHR (adjusted Hazard Ratio), CI (confidence interval)

### Per-protocol analysis of the primary outcome

The adjusted 2-year RD for the per-protocol effect of starting metformin monotherapy vs never starting any antidiabetic drug on COVID-19-related hospitalisation or death was 0.21% (95% CI - 0.08% to 0.51%). The adjusted RR was 1.11 (95% CI 0.96 to 1.29) and the adjusted HR was 1.11 (95% CI 0.96 to 1.29) with no substantial change in estimates when censoring weights were not truncated (appendix figure S5, appendix table S5).

### Intention-to-treat analysis of the secondary outcomes

The incidence of COVID-19 diagnosis was slightly higher in the metformin than the no antidiabetic group (maximally adjusted HR 1.07, 95% CI 1.01 to 1.15, figure 3). Within 2 years of follow-up, 33 (0.17%) of 19,599 participants in the metformin group had a recorded Long COVID diagnosis, compared to 33 (0.10%) of 32,739 participants in the no antidiabetic group (maximally adjusted HR 1.39, 95% CI 0.83 to 2.33, figure 3). Maximally adjusted HRs did not differ substantially from the age- and sex-adjusted HRs (appendix figure S3).

### Negative and positive control outcome analyses

When replacing the outcome of the primary model (Cox model, intention-to-treat) with bone fracture (negative control) and with diabetes-related deaths (positive control) the fully adjusted HRs were 1.00 (95% CI 0.81 to 1.22) and 1.41 (95% CI 1.09 to 1.81), respectively (figure 3).

### Subgroup analyses on the primary outcome

The subgroup analyses suggested some treatment effect heterogeneity for body mass index on COVID-19-related hospitalisation or death (appendix figure S6). There was no evidence for effect modification by any of the other prespecified subgroups (age, sex, ethnicity, deprivation, or HbA1c).

## Conclusions

Using a large, population-based cohort and new-user design, based on NHS electronic health records in England, this study estimated the effect of starting metformin within six months of T2D diagnosis on subsequent COVID-19 outcomes. Among more than 52,000 adults with newly diagnosed T2D, we found no protective effect of starting metformin, versus not starting, against COVID-19-related hospitalisation or death. Instead, after adjustment for an extensive set of confounders, starting metformin was associated with a slightly higher risk of COVID-19-related hospitalisation or death and COVID diagnoses. Results for the positive control outcome analysis suggested residual confounding, although the null result for the negative control outcome (bone fracture) was reassuring. Overall, the findings argue against a clinically meaningful benefit of starting metformin for prevention of severe or long COVID among people with T2D. Recorded Long COVID diagnoses were rare, and estimates imprecise.

Observational studies - and meta-analyses thereof - have consistently reported lower mortality or reduced risk of severe COVID-19 among metformin users compared with non-users.^5–8,15–17^ However, many of these studies were restricted to hospitalized populations and relied on prevalent-user designs, comparing long-term metformin users with heterogeneous control groups. Randomised trials, by contrast, have primarily evaluated starting metformin during the acute phase of COVID-19 in a broader at-risk population.^18,19^ A meta-analysis of these trials, as well as a large trial published after the meta-analysis, concluded that metformin has little to no effect on mortality and a small but uncertain effect on severe COVID-19 outcomes.^4,19^ Several factors may explain the discrepancy between observational and randomised evidence. First, T2D itself is a strong risk factor for severe COVID-19, and improved glycaemic control achieved with metformin may influence disease course independently of any direct antiviral or anti-inflammatory effects.^20^ While randomised trials largely excluded people with T2D - since they were already taking metformin and randomising them to metformin or placebo was unethical - observational studies were conducted mainly in the T2D population to satisfy the positivity assumption. Second, observational studies predominantly included hospitalized patients, while randomised trials recruited individuals in the community with non-severe COVID-19. Third, prevalent-user designs address the contrast of “being on metformin versus not being on metformin” at a given time, rather than the causal effect of starting metformin, which is the estimand targeted in randomised trials. In this respect, the present observational study more closely emulates the trial setting, as participants were drawn from primary care and a landmark new-user design was used to estimate the effect of *starting* metformin. The convergence of our results with randomised evidence underscores how design choices may explain some of the protective association seen in earlier observational work. More broadly, this illustrates how new-user designs can help reconcile discrepant observational and trial findings, and should be prioritised in future observational drug-repurposing studies.

The currently available evidence on metformin and Long COVID suggests a potential protective effect.^9^ Two randomised trials demonstrated that metformin started shortly after acute SARS-CoV-2 infection reduced incident Long COVID.^18,21^ A large observational analysis based on electronic health records from the US reported that prevalent use of metformin is associated with a reduced risk of Long COVID and an analysis based on English electronic health records - explicitly studying the starting of metformin after COVID-19 diagnosis - reported similar results.^22^ In contrast to the previous UK study, we restricted our Long COVID outcome to coded diagnoses only - which was likely underreporting the scale of Long COVID^14^, and our estimates were too imprecise to draw meaningful conclusions.

This study has several limitations. First, the landmark design introduces some concerns. Date of eligibility (T2D diagnosis) and start of follow-up are misaligned. In our specific case, however, selection bias at the landmark date due to differential outcome occurrence is impossible because the treatment allocation window closed before the pandemic began in the UK. There may still be some residual selection bias due to depletion of susceptibles with regards to all-cause death, though only 1377 (2.6%) deaths and loss to follow-up occurred between T2D diagnosis and landmark date with similar characteristics between individuals who were excluded and those who remained in the analysis (appendix table S3). Furthermore, estimates from this analysis have to be interpreted as conditional on still being alive at the time of reaching the six-month landmark. Second, underreporting and misclassification of the outcomes (particularly Long COVID) is likely. Possible misclassification of cause of death on death certificates cannot be excluded. Misclassification of exposure is also possible since we only observed metformin prescriptions but not dispensing or intake. Third, despite carefully defining a suitable study population with an indication for metformin (new T2D diagnosis with no contraindication to metformin, no extremely high HbA1c that would violate the positivity assumption) and adjusting for important confounders, unmeasured confounding is likely present, as suggested by the positive control outcome. The bimodal propensity score distribution implies that the untreated group includes a substantial proportion of people with genuinely mild dysglycaemia who may be healthier in ways not fully captured by measured confounders, which is the most plausible explanation for the hazard ratio above one. However, there was still reasonable propensity score overlap between the treatment groups, and estimates by HbA1c subgroups showed similar results. A further source of residual confounding is changes in covariates (e.g. HbA1c) between T2D diagnosis and the landmark date, when treatment status was assigned, because our models adjusted only for baseline values at T2D diagnosis. Finally, our results apply only to the T2D population and may not generalise to other populations.

In summary, this study aligns more closely with randomised evidence than with earlier observational reports, reinforcing the conclusion that starting metformin should not be expected to reduce severe COVID-19 risk among people with newly diagnosed T2D.

## Data Availability

The data in the NHS England OpenSAFELY Analytics Service is drawn from General Practice data across England where Optum and TPP are the data processors. This study uses data from TPP only. Linked data from hospitals and death registrations are also available.
Access to the underlying identifiable and potentially re-identifiable pseudonymised electronic health record data is tightly governed by various legislative and regulatory frameworks, and restricted by best practice. TPP developers initiate an automated process to create pseudonymised records in the core OpenSAFELY database, which are copies of key structured data tables in the identifiable records. These pseudonymised records are linked onto key external data resources that have also been pseudonymised via SHA-512 one-way hashing of NHS numbers using a shared salt. University of Oxford, Bennett Institute for Applied Data Science developers and PIs, who hold contracts with NHS England, have access to the OpenSAFELY pseudonymised data tables to develop the OpenSAFELY tools.
These tools in turn enable researchers with OpenSAFELY data access agreements to write and execute code for data management and data analysis without direct access to the underlying raw pseudonymised patient data, and to review the outputs of this code. All OpenSAFELY platform software is shared for inspection on GitHub: https://github.com/OpensAFELY-Core.
All code for the full data management pipeline, from raw data to completed results, for this analysis are available for review and reuse for all OpenSAFELY analyses (https://github.com/opensafely/metformin_covid).

## Administrative

## Acknowledgments

We acknowledge all the support received from the TPP Technical Operations team throughout this work, and for generous assistance from the information governance and database teams at NHS England and the NHS England Transformation Directorate. We acknowledge members of the OpenSAFELY Collaborative, who have contributed to the development and maintenance of the OpenSAFELY platform. We specifically want to thank Iain Dillingham (The Bennett Institute for Applied Data Science, University of Oxford) for his support with the diabetes algorithm reusable action (https://actions.opensafely.org/actions/diabetes-algo/v0.0.14/), which is a central part of this work.

## Information governance and ethical approval

This work was conducted with the approval of NHS England [project #175; https://www.opensafely.org/project/175/], and approved for publication following review by the Bennett Institute and the Department for Health and Social Care.

NHS England is the data controller of the NHS England OpenSAFELY Analytics Service; TPP is the data processor; all study authors using OpenSAFELY have the approval of NHS England.^23^ This implementation of OpenSAFELY is hosted within the TPP environment which is accredited to the ISO 27001 information security standard and is NHS IG Data Security and Information Governance Toolkit compliant.^24^ Patient data has been pseudonymised for analysis and linkage using industry standard cryptographic hashing techniques; all pseudonymised datasets transmitted for linkage onto OpenSAFELY are encrypted; access to the NHS England OpenSAFELY Analytics Service is via a virtual private network (VPN) connection; the researchers hold contracts with NHS England and only access the platform to initiate database queries and statistical models; all database activity is logged; only aggregate statistical outputs leave the platform environment following best practice for anonymisation of results such as statistical disclosure control for low cell counts.^25^ The service adheres to the obligations of the UK General Data Protection Regulation (UK GDPR) and the Data Protection Act 2018. The Secretary of State has requested that NHS England operates the Service under the COVID-19 Directions 2020^26^ and the NHS OpenSAFELY Data Analytics Service Pilot Directions 2025 which came into force on 17 June 2025.^27^ The Pilot Directions enable the Service to be operated for purposes wider than COVID-19. Taken together, these provide the legal bases to link patient datasets using the service.

## Funding

The OpenSAFELY platform is principally funded by grants from NHS England [2023-2026], the Wellcome Trust (222097/Z/20/Z [2020-2024] and 311535/Z/24/Z [2025-2031]), the Medical Research Council (MRC) (MR/V015737/1 [2020-2021]). Additional contributions to OpenSAFELY have been funded by grants from Medical Research Council (MRC) via the National Core Study programme Longitudinal Health and Wellbeing strand (MC_PC_20030, MC_PC_20059 [2020-2022]) and the Data and Connectivity strand (MC_PC_20058 [2021-2022]), the National Institute for Health Research (NIHR) and the Medical Research Council (MRC) via the CONVALESCENCE programme (COV-LT-0009, MC_PC_20051 [2021-2024]), NHS England via the Primary Care Medicines Analytics Unit [2021-2024]. This study was also supported by the European Union’s Horizon Europe research and innovation programme Grant Agreement No. 101156304 for PROACT EU RESPONSE. AA receives his salary from the postdoc mobility return grant of the Swiss National Science Foundation (#P5R5-3_239044). VW is supported by the UK Medical Research Council Integrative Epidemiology Unit (MC_UU_00032/3). The views expressed are those of the authors and not necessarily those of the NIHR, Wellcome Trust, NHS England, Wellcome Trust, the Department of Health and Social Care, Horizon or other funders. Funders had no role in the study design, collection, analysis, and interpretation of data; in the writing of the report; and in the decision to submit the article for publication.

## Contributors

Conceptualisation: AA, VW, JACS; Methodology: AA, VW, PMD, RD, MZ, TP, SVE, WJH, JACS; Formal analysis: AA; Writing-original draft: AA; Writing-review & editing: AA, VW, PMD, RD, MZ, TP, SVE, WJH, JACS; Visualisation: AA; Project administration: AA, VW, RD, JACS; Funding acquisition: AA, VW, WJH, JACS. All authors had access to the aggregated data included in the manuscript. All authors contributed to and approved the final manuscript.

## Declaration of competing interests

The authors have declared no competing interest.

## Patient and public involvement and engagement

OpenSAFELY has involved patients and the public in various ways: we developed a public website that provides a detailed description of the platform in language suitable for a lay audience (https://opensafely.org); we have participated in two citizen juries exploring public trust in OpenSAFELY; we have co-developed an explainer video (https://www.opensafely.org/about/); we have patient representation who are experts by experience on our OpenSAFELY Oversight Board; we have partnered with Understanding Patient Data to produce lay explainers on the importance of large datasets for research; we have presented at various online public engagement events to key communities (e.g., Healthcare Excellence Through Technology; Faculty of Clinical Informatics annual conference; NHS Assembly; HDRUK symposium); and more. To ensure the patient voice is represented, we are working closely to decide on language choices with appropriate medical research charities (e.g., Association of Medical Research Charities). We will share information and interpretation of our findings through press releases, social media channels, and plain language summaries.

## Data access and verification

The data in the NHS England OpenSAFELY Analytics Service is drawn from General Practice data across England where Optum and TPP are the data processors. This study uses data from TPP only. Linked data from hospitals and death registrations are also available.

Access to the underlying identifiable and potentially re-identifiable pseudonymised electronic health record data is tightly governed by various legislative and regulatory frameworks, and restricted by best practice. TPP developers initiate an automated process to create pseudonymised records in the core OpenSAFELY database, which are copies of key structured data tables in the identifiable records.

These pseudonymised records are linked onto key external data resources that have also been pseudonymised via SHA-512 one-way hashing of NHS numbers using a shared salt. University of Oxford, Bennett Institute for Applied Data Science developers and PIs, who hold contracts with NHS England, have access to the OpenSAFELY pseudonymised data tables to develop the OpenSAFELY tools.

These tools in turn enable researchers with OpenSAFELY data access agreements to write and execute code for data management and data analysis without direct access to the underlying raw pseudonymised patient data, and to review the outputs of this code. All OpenSAFELY platform software is shared for inspection on GitHub: https://github.com/OpensAFELY-Core.

All code for the full data management pipeline, from raw data to completed results, for this analysis are available for review and reuse for all OpenSAFELY analyses (https://github.com/opensafely/metformin_covid). The data management and analysis code for this paper was led by AA and contributed to by VW, PMD, TP, and WJH.

## Notes

### Clinical Protocols

https://github.com/opensafely/metformin_covid/tree/main/protocol

### Author Declarations

This work was conducted with the approval of NHS England [project #175; https://www.opensafely.org/project/175/], and approved for publication following review by the Bennett Institute and the Department for Health and Social Care. NHS England is the data controller of the NHS England OpenSAFELY Analytics Service; TPP is the data processor; all study authors using OpenSAFELY have the approval of NHS England. This implementation of OpenSAFELY is hosted within the TPP environment which is accredited to the ISO 27001 information security standard and is NHS IG Data Security and Information Governance Toolkit compliant. Patient data has been pseudonymised for analysis and linkage using industry standard cryptographic hashing techniques; all pseudonymised datasets transmitted for linkage onto OpenSAFELY are encrypted; access to the NHS England OpenSAFELY Analytics Service is via a virtual private network (VPN) connection; the researchers hold contracts with NHS England and only access the platform to initiate database queries and statistical models; all database activity is logged; only aggregate statistical outputs leave the platform environment following best practice for anonymisation of results such as statistical disclosure control for low cell counts. The service adheres to the obligations of the UK General Data Protection Regulation (UK GDPR) and the Data Protection Act 2018. The Secretary of State has requested that NHS England operates the Service under the COVID-19 Directions 202026 and the NHS OpenSAFELY Data Analytics Service Pilot Directions 2025 which came into force on 17 June 2025. The Pilot Directions enable the Service to be operated for purposes wider than COVID-19. Taken together, these provide the legal bases to link patient datasets using the service.

## References

1. Siedner MJ, Sax PE. Repurposing Revisited: Exploring the Role of Metformin for Treatment of COVID-19. Clin Infect Dis. 2024 Aug 15;79(2):292–4. doi:10.1093/cid/ciae154

2. Samuel SM, Varghese E, Büsselberg D. Therapeutic Potential of Metformin in COVID-19: Reasoning for Its Protective Role. Trends Microbiol. 2021 Oct 1;29(10):894–907. doi:10.1016/j.tim.2021.03.004 PubMed PMID: 33785249.

3. Bramante CT, Beckman KB, Mehta T, Karger AB, Odde DJ, Tignanelli CJ, et al. Favorable Antiviral Effect of Metformin on SARS-CoV-2 Viral Load in a Randomized, Placebo-Controlled Clinical Trial of COVID-19. Clin Infect Dis. 2024 Aug 15;79(2):354–63. doi:10.1093/cid/ciae159

4. Chowdhury SR, Islam N, Zhou Q, Hasan MK, Chowdhury MR, Siemieniuk RA, et al. Metformin for covid-19: systematic review and meta-analysis of randomised controlled trials. BMJ Med. 2025 May 21;4(1). doi:10.1136/bmjmed-2024-001126 PubMed PMID: 10.1136/bmjmed-2024-001126.

5. Zhan K, Weng L, Qi L, Wang L, Lin H, Fang X, et al. Effect of Antidiabetic Therapy on Clinical Outcomes of COVID-19 Patients With Type 2 Diabetes: A Systematic Review and Meta-Analysis. Ann Pharmacother. 2023 Jul;57(7):776–86. doi:10.1177/10600280221133577 PubMed PMID: 36314281; PubMed Central PMCID: PMC9618918.

6. Song ZH, Huang QM, Xu SS, Zhou JB, Zhang C. The Effect of Antihyperglycemic Medications on COVID-19: A Meta-analysis and Systematic Review from Observational Studies. Ther Innov Regul Sci. 2024 Jul;58(4):773–87. doi:10.1007/s43441-024-00633-6 PubMed PMID: 38683419.

7. Keels JN, McDonald IR, Lee CS, Dwyer AA. Antidiabetic agent use and clinical outcomes in patients with diabetes hospitalized for COVID-19: a systematic review and meta-analysis. Front Endocrinol. 2024;15:1482853. doi:10.3389/fendo.2024.1482853 PubMed PMID: 39835258; PubMed Central PMCID: PMC11743176.

8. Petrelli F, Grappasonni I, Nguyen CTT, Tesauro M, Pantanetti P, Xhafa S, et al. Metformin and Covid-19: a systematic review of systematic reviews with meta-analysis. Acta Bio-Medica Atenei Parm. 2023 Aug 30;94(S3):e2023138. doi:10.23750/abm.v94iS3.14405 PubMed PMID: 37695186.

9. Bramante CT, Boulware DR. Preventing Long COVID With Metformin. Clin Infect Dis. 2026 Mar 17;82(3):e433–5. doi:10.1093/cid/ciaf700

10. Nab L, Schaffer AL, Hulme W, DeVito NJ, Dillingham I, Wiedemann M, et al. OpenSAFELY: A platform for analysing electronic health records designed for reproducible research. Pharmacoepidemiol Drug Saf. 2024;33(6):e5815. doi:10.1002/pds.5815

11. Hernán MA, Sterne JAC, Higgins JPT, Shrier I, Hernández-Díaz S. A Structural Description of Biases That Generate Immortal Time. Epidemiol Camb Mass. 2025 Jan 1;36(1):107–14. doi:10.1097/EDE.0000000000001808 PubMed PMID: 39494894; PubMed Central PMCID: PMC11598638.

12. Langan SM, Schmidt SA, Wing K, Ehrenstein V, Nicholls SG, Filion KB, et al. The reporting of studies conducted using observational routinely collected health data statement for pharmacoepidemiology (RECORD-PE). BMJ. 2018 Nov 14;363:k3532. doi:10.1136/bmj.k3532 PubMed PMID: 30429167.

13. Thygesen JH, Tomlinson C, Hollings S, Mizani MA, Handy A, Akbari A, et al. COVID-19 trajectories among 57 million adults in England: a cohort study using electronic health records. Lancet Digit Health. 2022 Jul 1;4(7):e542–57. doi:10.1016/S2589-7500(22)00091-7 PubMed PMID: 35690576.

14. Henderson AD, Butler-Cole BF, Tazare J, Tomlinson LA, Marks M, Jit M, et al. Clinical coding of long COVID in primary care 2020-2023 in a cohort of 19 million adults: an OpenSAFELY analysis. EClinicalMedicine. 2024 Jun;72:102638. doi:10.1016/j.eclinm.2024.102638 PubMed PMID: 38800803; PubMed Central PMCID: PMC11127160.

15. Khunti K, Knighton P, Zaccardi F, Bakhai C, Barron E, Holman N, et al. Prescription of glucose-lowering therapies and risk of COVID-19 mortality in people with type 2 diabetes: a nationwide observational study in England. Lancet Diabetes Endocrinol. 2021 May;9(5):293–303. doi:10.1016/S2213-8587(21)00050-4

16. Oh TK, Song IA. Metformin use and risk of COVID-19 among patients with type II diabetes mellitus: an NHIS-COVID-19 database cohort study. Acta Diabetol. 2021 Jun;58(6):771–8. doi:10.1007/s00592-020-01666-7 PubMed PMID: 33582839; PubMed Central PMCID: PMC7882044.

17. Ojeda-Fernández L, Foresta A, Macaluso G, Colacioppo P, Tettamanti M, Zambon A, et al. Metformin use is associated with a decrease in the risk of hospitalization and mortality in COVID -19 patients with diabetes: A population-based study in Lombardy. Diabetes Obes Metab. 2022 May;24(5):891–8. doi:10.1111/dom.14648

18. Bramante CT, Stewart TG, Boulware DR, McCarthy MW, Gao Y, Rothman RL, et al. Metformin on the Presence of COVID-19 Symptoms 6 Months After Infection: The ACTIV-6 Randomized Clinical Trial. Clin Infect Dis. 2026 Jun 4;ciag335. doi:10.1093/cid/ciag335

19. Bramante CT, Stewart TG, Boulware DR, McCarthy MW, Gao Y, Rothman RL, et al. Metformin and Time to Sustained Recovery in Adults With COVID-19: The ACTIV-6 Randomized Clinical Trial. JAMA Intern Med. 2025 Sep 1;185(9):1092–101. doi:10.1001/jamainternmed.2025.2570 PubMed PMID: 40658388; PubMed Central PMCID: PMC12261116.

20. Apicella M, Campopiano MC, Mantuano M, Mazoni L, Coppelli A, Del Prato S. COVID-19 in people with diabetes: understanding the reasons for worse outcomes. Lancet Diabetes Endocrinol. 2020 Sep;8(9):782–92. doi:10.1016/S2213-8587(20)30238-2

21. Bramante CT, Buse JB, Liebovitz DM, Nicklas JM, Puskarich MA, Cohen K, et al. Outpatient treatment of COVID-19 and incidence of post-COVID-19 condition over 10 months (COVID-OUT): a multicentre, randomised, quadruple-blind, parallel-group, phase 3 trial. Lancet Infect Dis. 2023 Oct 1;23(10):1119–29. doi:10.1016/S1473-3099(23)00299-2 PubMed PMID: 37302406.

22. Chaichana U, Man KKC, Ju C, Makaronidis J, Wei L. Effect of Metformin on the Risk of Post-COVID-19 Condition Among Individuals With Overweight or Obese: A Population-based Retrospective Cohort Study. Clin Infect Dis. 2026 Mar 17;82(3):e423–32. doi:10.1093/cid/ciaf429

23. NHS England Digital [Internet]. [cited 2025 Apr 30]. The NHS England OpenSAFELY COVID-19 service - privacy notice. Available from: https://digital.nhs.uk/coronavirus/coronavirus-covid-19-response-information-governance-hub/the-nhs-england-opensafely-covid-19-service-privacy-notice

24. Data Security and Protection Toolkit - NHS England Digital [Internet]. [cited 2025 Aug 1]. Available from: https://web.archive.org/web/20250405100349/ https://digital.nhs.uk/services/data-security-and-protection-toolkit/data-security-and-protection-toolkit

25. NHS England Digital [Internet]. [cited 2025 Aug 1]. ISB1523: Anonymisation Standard for Publishing Health and Social Care Data. Available from: https://digital.nhs.uk/data-and-information/information-standards/governance/latest-activity/standards-and-collections/isb1523-anonymisation-standard-for-publishing-health-and-social-care-data

26. NHS England Digital [Internet]. [cited 2025 Aug 1]. COVID-19 Public Health Directions 2020. Available from: https://digital.nhs.uk/about-nhs-digital/corporate-information-and-documents/directions-and-data-provision-notices/secretary-of-state-directions/covid-19-public-health-directions-2020

27. NHS England Digital [Internet]. [cited 2025 Jul 13]. NHS OpenSAFELY Data Analytics Service Pilot Directions 2025. Available from: https://digital.nhs.uk/about-nhs-digital/corporate-information-and-documents/directions-and-data-provision-notices/secretary-of-state-directions/nhs-opensafely-data-analytics-service-pilot-directions-2025

