## Appendix for "Effect of starting metformin on severe and long COVID-19 among people living with type 2 diabetes in England: a new user cohort study in OpenSAFELY-TPP"

**APPENDIX** ... 1

**TEXT** ... 2

**Appendix Text S1:** Details regarding eligibility and quality assurance variables ... 2

**Appendix Text S2:** RECORD-PE checklist ... 6

**Appendix Text S3:** Confounders/Covariates ... 11

**TABLES** ... 17

**Appendix Table S1:** Exposure/treatment variables ... 17

**Appendix Table S2:** Outcome variables ... 18

**Appendix Table S3:** Characteristics of people who died or loss to follow-up until landmark ... 19

**Appendix Table S4:** Characteristics of people who died or loss to follow-up until pandemic start ... 21

**Appendix Table S5.** 2-year risk estimates for the primary outcome ... 23

**FIGURES** ... 24

**Appendix Figure S1:** Directed acyclic graph ... 24

**Appendix Figure S2.** Density plots (smoothed histogram), based on propensity score of receiving metformin ... 25

**Appendix Figure S3.** All outcomes, minimal adjusted model, primary analysis (Cox model, intention-to-treat) ... 26

**Appendix Figure S4.** Primary outcome, cumulative incidence curves, weighted Kaplan Meier (intention-to-treat) ... 27

**Appendix Figure S5.** Primary outcome, cumulative incidence curves, pooled logistic regression (per-protocol) ... 28

**Appendix Figure S6.** Forest plot, presenting subgroup analyses and tests for heterogeneity (Cochran's Q) on primary outcome ... 29

### TEXT

**Appendix Text S1:** Details regarding eligibility and quality assurance variables

Inclusion Criteria:

1) Aged ≥ 18 years & ≤ 85 years
2) Diagnosis of T2DM between 01.08.2018 to 01.08.2019
3) Registered for at least a year with a GP in England who uses TPP software

Exclusion Criteria:

1) Hospitalized, defined as having an admission to hospital record but no discharge record at baseline (i.e. T2D diagnosis)
2) In a care home, defined as having a record of long-stay nursing/care home at baseline
3) In palliative care, defined as having a record of palliative care within 6 months prior to baseline
4) Any use of metformin or another major glucose-lowering medication (SGLT2, DPP4, sulfonylurea, thiazolidinedione, GLP-1, meglitinides, alpha-glucosidase-inhibitors, insulin) prior to baseline
5) Recorded HbA1c, within 2 years prior to baseline
6) Most recent (within 2 years prior to baseline) HbA1c is above 75 mmol/mol
7) Known hypersensitivity / intolerance to metformin
8) Any history of moderate to severe renal impairment (eGFR of <30ml/min/1.73 m2; chronic kidney diseases stage 4/5)
9) Any history of advanced decompensated liver cirrhosis
10) Use of the following medications in the past 14 days: Cimetidine, hydroxychloroquine, tafenoquine, patiromer, ranolazine, Monoamine Oxide Inhibitors (Phenelzine, Tranylcypromine, Selegiline, Isocarboxazide, moclobemide), Alpha-1 antagonists, Sotalol, Clonidine, Phosphodiesterase 5 inhibitors, Methyldopa, Prazosin, Terasozin, Doxazosin)

Details about each eligibility variable, their data sources, lookback period, and codelists are available in table (a) below.

In addition, we apply the following data quality assurance and data completeness criteria:
Data quality assurance criteria:

1) Remove individuals whose year of birth is missing
2) Remove individuals whose year of birth is after their year of death
3) Remove individuals whose year of birth is after the study end date or who were older than 110 years when the UK National Health Services (NHS) was established (1838, since NHS was established in 1948)
4) Remove individuals whose date of death is on or before 01/01/1900 or after current date (Sys.Date())
5) Remove men whose records contain pregnancy codes
6) Remove men whose records contain hormone replacement therapy or combined contraceptive pill medication codes
7) Remove women whose records contain prostate cancer codes

Data completeness criteria:
Patients will be included only if they meet all completeness criteria:

1) Known sex (female/male)
2) Known deprivation (Index of Multiple Deprivation [IMD]) at baseline date
3) Known sustainability and transformation partnership (STP) region, an NHS administrative geographical area, at baseline

Details regarding data quality and data completeness criteria variables are presented in table (b) below.

**Table (a) Eligibility variables**

| Variable | Name | Type | Definition | Data sources (*tpp schema* in brackets) | Notes |
|---|---|---|---|---|---|
| New type 2 diabetes mellitus (T2DM) diagnosis | elig_date_t2dm | Date | Date | 1) Primary care: Diagnoses *(clinical_events):*<br>- diabetes_type1_ctv3_clinical<br>- diabetes_type2_ctv3_clinical<br>- diabetes_other_ctv3_clinical<br>- diabetes_gestational_ctv3_clinical<br>- diabetes_diagnostic_ctv3_clinical<br><br>2) Primary care: Drugs *(medications):*<br>- insulin_dmd<br>- antidiabetic_drugs_snomed_clinical<br>- non_metformin_dmd<br><br>3) Secondary care: Diagnoses *(apcs):*<br>- diabetes_type1_icd10<br>- diabetes_type2_icd10<br>- diabetes_gestational_icd10 | In ehrQL: search for **first code** before studyend_date, apply diagnosis window later in R<br><br>See Appendix 1 regarding the algorithm to define Diabetes types.<br><br>**The final diabetes variable (elig_date_t2dm) is extracted through the adjudication algorithm (reusable action): https://www.bennett.ox.ac.uk/blog/2025/05/introducing-diabetes-algo-a-reusable-action-for-opensafely-users/.** |
| Any metformin use, before baseline | elig_date_metfin_first (= exp_date_metfin_first) | Date | Date | Primary care (medications):<br>- metformin_dmd | In ehrQL: search for **first code** before elig_date_t2dm |
| Any other antidiabetic use, before baseline | elig_date_***_first | Date | Date | Primary care (medications)***:<br>Individual codelists for each medication see in appendix Table S1 (intervention/exposure) | In ehrQL: search for **first code** before elig_date_t2dm |
| Known hypersensitivity and / or intolerance to metformin, before baseline | elig_date_metfin_allergy_first | Date | Date | Primary care (clinical_events):<br>- metformin_allergy_snomed_clinical | In ehrQL: search for **first code** before elig_date_t2dm |
| Clinical history of moderate to severe renal impairment (eGFR of <30ml/min/1.73 m2; chronic stage 4/5), before baseline | elig_date_ckd_45_first | Date | Date | 1) Primary care (clinical_events):<br>- ckd_snomed_clinical_45<br>2) HES APC (apcs):<br>- ckd_stage4_icd10 -> ["N184"]<br>- ckd_stage5_icd10 -> ["N185"] | In ehrQL: search for **first code** before elig_date_t2dm |

| | | | | | |
|---|---|---|---|---|---|
| Clinical history of advance decompensated liver cirrhosis, before baseline | elig_date_liver_cirrhosis_first | Date | Date | 1) Primary care (clinical_events):<br>- advanced_decompensated_cirrhosis_snomed_clinical<br>- ascitic_drainage_snomed_clinical<br>2) HES APC (apcs):<br>- advanced_decompensated_cirrhosis_icd10 | In ehrQL: search for **first code** before elig_date_t2dm |
| Use of the following medications in the last 14 days: Cimetidine, hydroxychloroquine, dolutegravir, patiromer, ranolazine, Monoamine Oxide Inhibitors (Phenelzine, Tranylcypromine, Selegiline, moclobemide), Sotalol, Clonidine, Methyldopa, Prazosin, Doxazosin, before baseline | elig_date_metfin_interaction_last | Date | Date | Primary care (medications):<br>- metformin_interaction_dmd | In ehrQL: search for **first code** before elig_date_t2dm |
| Hospitalized, within 2 days prior to baseline | elig_bin_hosp | Binary | T, F | HES APC (apcs):<br>- any admission date within 2 days prior to baseline | Baseline: elig_date_t2dm |
| Care home resident, at baseline | elig_bin_carehome_status | Binary | T, F | 1) Primary care (clinical_events):<br>- carehome<br>2) Primary care (addresses):<br>- care_home_is_potential_match<br>- care_home_requires_nursing<br>- care_home_does_not_require_nursing | Baseline: elig_date_t2dm |
| Palliative care, within 6 months prior to baseline | elig_date_palliative | Date | Date | 1) Primary care (clinical_events):<br>- palliative_snomed | Baseline: elig_date_t2dm |

**Table (b) Data completeness and quality assurance variables**

| Variable | Name | Type | Definition | Data sources (tpp schema in brackets) | Notes |
|---|---|---|---|---|---|
| Known sex | qa_bin_is_female_or_male | Binary | T, F | Primary care (patients) | Based on entire EHR |

| | | | | | |
|---|---|---|---|---|---|
| Known age between 18 and 110 inclusive | qa_bin_was_adult | Binary | T, F | Primary care (patients) | Based elig_date_t2dm |
| Alive | qa_bin_was_alive | Binary | T, F | Primary care (patients) | Based elig_date_t2dm |
| Known deprivation (IMD) | qa_bin_known_imd | Binary | T, F | Primary care (practice_registration) | Based elig_date_t2dm |
| Known region | *based on same variable as below* | | | | |
| Registered in an English GP with TPP for at least past 12 months | qa_bin_was_registered | Binary | T, F | Primary care (practice_registration) | 1 year = 365.25 days, taking into account leap years. Based elig_date_t2dm |
| Year of Birth | qa_num_birth_year | Numeric | continuous (year only) | Primary care (patients) | Based on entire EHR |
| Date of Death | qa_date_of_death | Date | Date | Death certificates (ons_deaths) | Based on entire EHR |
| Pregnancy | qa_bin_pregnancy | Binary | T, F | Primary care (clinical_events): pregnancy_snomed_clinical | Based on entire EHR, |
| Combined oral contraceptive pill | qa_bin_cocp | Binary | T, F | Primary care (medications): cocp_dmd | Based on entire EHR |
| Hormone replacement therapy | qa_bin_hrt | Binary | T, F | Primary care (medications): hrt_dmd | Based on entire EHR |
| Prostata cancer diagnosis | qa_bin_prostate_cance r | Binary | T, F | 1) Primary care (clinical_events): prostate_cancer_snomed_clinical<br>3) HES APC (apcs): prostate_cancer_icd10<br>3) Death certificates (ons_deaths): prostate_cancer_icd10 | Based on entire EHR<br><br>Re ons_deaths: Search in underlying_cause_of_death as well as in all other 15 cause_of_death fields |

**Appendix Text S2:** RECORD-PE checklist

| Item No | STROBE items | RECORD items | RECORD-PE items | Page No |
|---|---|---|---|---|
| **Title and abstract** | | | | |
| 1 | (a) Indicate the study's design with a commonly used term in the title or the abstract.<br>(b) Provide in the abstract an informative and balanced summary of what was done and what was found. | 1.1: The type of data used should be specified in the title or abstract. When possible, the name of the databases used should be included. 1.2: If applicable, the geographical region and timeframe within which the study took place should be reported in the title or abstract. 1.3: If linkage between databases was conducted for the study, this should be clearly stated in the title or abstract. | — | Page 2 |
| **Introduction** | | | | |
| Background rationale | | | | |
| 2 | Explain the scientific background and rationale for the investigation being reported. | — | — | Page 4 |
| Objectives | | | | |
| 3 | State specific objectives, including any prespecified hypotheses. | — | — | Page 4 |
| **Methods** | | | | |
| Study design | | | | |
| 4 | Present key elements of study design early in the paper. | — | 4.a: Include details of the specific study design (and its features) and report the use of multiple designs if used.<br>4.b: The use of a diagram(s) is recommended to illustrate key aspects of the study design(s), including exposure, washout, lag and observation periods, and covariate definitions as relevant. | Page 5, figure 1 |
| Setting | | | | |
| 5 | Describe the setting, locations, and relevant dates, including periods of recruitment, exposure, follow-up, and data collection. | — | — | Page 5 and 6 |
| Participants | | | | |

| | | | | |
|---|---|---|---|---|
| 6 | (a) Cohort study—give the eligibility criteria, and the sources and methods of selection of participants. Describe methods of follow-up. Case-control study—give the eligibility criteria, and the sources and methods of case ascertainment and control selection. Give the rationale for the choice of cases and controls. Cross sectional study—give the eligibility criteria, and the sources and methods of selection of participants.<br>(b) Cohort study—for matched studies, give matching criteria and number of exposed and unexposed. Case-control study—for matched studies, give matching | 6.1: The methods of study population selection (such as codes or algorithms used to identify participants) should be listed in detail. If this is not possible, an explanation should be provided. 6.2: Any validation studies of the codes or algorithms used to select the population should be referenced. If validation was conducted for this study and not published elsewhere, detailed methods and results should be provided.<br>6.3: If the study involved linkage of databases, consider use of a flow | 6.1.a: Describe the study entry criteria and the order in which these criteria were applied to identify the study population. Specify whether only users with a specific indication were included and whether patients were allowed to enter the study population once or if multiple entries were permitted. See explanatory document for guidance related to matched designs. | Page 5 |
| | criteria and the number of controls per case. | diagram or other graphical display to demonstrate the data linkage process, including the number of individuals with linked data at each stage. | | Page 4 |
| Variables | | | | |
| 7 | Clearly define all outcomes, exposures, predictors, potential confounders, and effect modifiers. Give diagnostic criteria, if applicable. | 7.1: A complete list of codes and algorithms used to classify exposures, outcomes, confounders, and effect modifiers should be provided. If these cannot be reported, an explanation should be provided. | 7.1.a: Describe how the drug exposure definition was developed.<br>7.1.b: Specify the data sources from which drug exposure information for individuals was obtained.<br>7.1.c: Describe the time window(s) during which an individual is considered exposed to the drug(s). The rationale for selecting a particular time window should be provided. The extent of potential left truncation or left censoring should be specified.<br>7.1.d: Justify how events are attributed to current, prior, ever, or cumulative drug exposure.<br>7.1.e: When examining drug dose and risk attribution, describe how current, historical or time on therapy are considered.<br>7.1.f: Use of any comparator groups should be outlined and justified.<br>7.1.g: Outline the approach used to handle individuals with more than one relevant drug exposure during the study period. | Page 5 and appendix table S1 |
| Data sources/measurement | | | | |
| 8 | For each variable of interest, give sources of data and details of methods of assessment (measurement). Describe comparability of assessment methods if there is more than one group. | — | 8.a: Describe the healthcare system and mechanisms for generating the drug exposure records. Specify the care setting in which the drug(s) of interest was prescribed. | Page 4a and section “Administrative” |
| Bias | | | | |
| 9 | Describe any efforts to address potential sources of bias. | — | — | Page 5 (study design) and page 7 (sensitivi |

| | | | | |
|---|---|---|---|---|
| | | | | ty analyses ) |
| Study size | | | | |
| 10 | Explain how the study size was arrived at. | — | — | Page 5 |
| Quantitative variables | | | | |
| 11 | Explain how quantitative variables were handled in the analyses. If applicable, describe which groupings were chosen, and why. | — | — | Page 6 and 7 and appendix text S3 (covariates) and appendix table S2 (outcomes) |
| Statistical methods | | | | |
| 12 | (a) Describe all statistical methods, including those used to control for confounding.<br>(b) Describe any methods used to examine subgroups and interactions.<br>(c) Explain how missing data were addressed.<br>(d) Cohort study—if applicable, explain how loss to follow-up was addressed. Case-control study—if applicable, explain how matching of cases and controls was addressed. Cross sectional study—if applicable, describe analytical methods taking account of sampling strategy. (e) Describe any sensitivity analyses. | — | 12.1.a: Describe the methods used to evaluate whether the assumptions have been met.<br>12.1.b: Describe and justify the use of multiple designs, design features, or analytical approaches. | Page 6 and 7 |
| Data access and cleaning methods | | | | |
| 12 | — | 12.1: Authors should describe the extent to which the investigators had access to the database population used to create the study population.<br>12.2: Authors should provide information on the data cleaning methods used in the study. | — | Section "Administrative" |
| Linkage | | | | |
| 12 | — | 12.3: State whether the study included person level, institutional level, or other data linkage across two or more databases. The methods of linkage and methods of linkage quality evaluation should be provided. | — | Page 4 (data sources) |
| **Results** | | | | |

| | | | | |
|---|---|---|---|---|
| Participants | | | | |
| 13 | (a) Report the numbers of individuals at each stage of the study (eg, numbers potentially eligible, examined for eligibility, confirmed eligible, included in the study, completing follow-up, and analysed).<br>(b) Give reasons for non-participation at each stage.<br>(c) Consider use of a flow diagram. | 13.1: Describe in detail the selection of the individuals included in the study (that is, study population selection) including filtering based on data quality, data availability, and linkage. The selection of included individuals can be described in the text or by means of the study flow diagram. | — | Page 7 (Results) and flow diagram figure 2 |
| Descriptive data | | | | |
| 14 | (a) Give characteristics of study participants (eg, demographic, clinical, social) and information on exposures and potential confounders.<br>(b) Indicate the number of participants with missing data for each variable of interest.<br>(c) Cohort study—summarise follow-up time (eg, average and total amount). | — | — | Page 7 |
| Outcome data | | | | |
| 15 | Cohort study—report numbers of outcome events or summary measures over time. Case-control study—report numbers in each exposure category, or summary measures of exposure. Cross sectional study—report numbers of outcome events or summary measures. | — | — | Page 7 and 8 |
| Main results | | | | |
| 16 | (a) Give unadjusted estimates and, if applicable, confounder adjusted estimates and their precision (eg, 95% confidence intervals). Make clear which confounders were adjusted for and why they were included.<br>(b) Report category boundaries when continuous variables are categorised. | — | — | Page 7 and 8 |

| | | | | |
|---|---|---|---|---|
| | (c) If relevant, consider translating estimates of relative risk into absolute risk for a meaningful time period. | | | Page 7 and 8 |
| Other analyses | | | | |
| 17 | Report other analyses done—eg, analyses of subgroups and interactions, and sensitivity analyses. | — | — | Page 7 and 8 |
| **Discussion** | | | | |
| Key results | | | | |
| 18 | Summarise key results with reference to study objectives. | — | — | Page 8 and 9 |
| Limitations | | | | |

| | | | | |
|---|---|---|---|---|
| 19 | Discuss limitations of the study, taking into account sources of potential bias or imprecision. Discuss both direction and magnitude of any potential bias. | 19.1: Discuss the implications of using data that were not created or collected to answer the specific research question(s). Include discussion of misclassification bias, unmeasured confounding, missing data, and changing eligibility over time, as they pertain to the study being reported. | 19.1.a: Describe the degree to which the chosen database(s) adequately captures the drug exposure(s) of interest. | Page 8 and 9 |
| Interpretation | | | | |
| 20 | Give a cautious overall interpretation of results considering objectives, limitations, multiplicity of analyses, results from similar studies, and other relevant evidence. | — | 20.a: Discuss the potential for confounding by indication, contraindication or disease severity or selection bias (healthy adherer/sick stopper) as alternative explanations for the study findings when relevant. | Page 8 and 9 |
| Generalisability | | | | |
| 21 | Discuss the generalisability (external validity) of the study results. | — | — | Page 8 and 9 |
| **Other information** | | | | |
| Funding | | | | |
| 22 | Give the source of funding and the role of the funders for the present study and, if applicable, for the original study on which the present article is based. | — | — | Section “Administrative” |
| Accessibility of protocol, raw data, and programming code | | | | |
| 22 | — | 22.1: Authors should provide information on how to access any supplemental information such as the study protocol, raw data, or programming code. | — | Section “Administrative” |

**Appendix Text S3:** Confounders/Covariates

The covariates, which were pre-specified as potentially important confounding variables are listed below, with further details in table (a) below. Unless otherwise specified, variables are created using diagnostic codes present ever in a patients' medical record and are defined at baseline (T2DM diagnosis):

- Age: splines with knot locations based on 10, 50 and 90 percentile
- Sex: binary (male, female)
- Ethnicity: categorical (White; Black or Black British; Asian or Asian British; Mixed or Unknown or Other)
- Calendar time of T2DM diagnosis: continuous
- Index of Multiple Deprivation ([IMD], derived as quintiles, from the patient's postcode at lower super output area level): categorical
- Rural/urban area: categorical (Urban conurbation; Urban city or town; Rural town or village)Health care worker occupation: logical
- Primary care consultation rate in previous year: continuous
- Smoking status: categorical (Current smoker; Never smoker; Ever or missing)
- BMI: categorical (BMI 30+ or obesity diagnosis; BMI 25-30; BMI below 25 or missing), most recent value in past 2 years
- HbA1c in mmol/mol: continuous, log-transformed), most recent value in past 2 years
- Number of HbA1c measurements in previous 2 years: continuous
- Diabetes-related complications: logical (diabetic foot, nephropathy, neuropathy, or retinopathy)
- Total cholesterol/high-density lipoprotein cholesterol ratio: categorical (below 3.5:1; 3.5:1 to 5:1; above 5:1; missing), most recent value in past 2 years
- Prediabetes: logical (clinical code or HbA1c in prediabetes range [42-47.9 mmol/mol])
- Comorbidities: logical (acute myocardial infarction, stroke, other arterial embolism, venous thromboembolism, heart failure, angina, dementia, cancer, hypertension, depression, chronic obstructive pulmonary disease, liver disease, chronic kidney disease, or polycystic ovary syndrome)
- Region as stratification factor: categorical (7 regions)

All comorbidities are identified through SNOMED CT codes in primary care records and ICD-10 in secondary care records. In case of convergence or disclosure risk, we may collapse categories further, e.g. for ethnicity or smoking status.

##### Table (a). Demographic variables, covariates and potential confounders

| Variable | Name | Type | Definition | Data sources | Notes |
|---|---|---|---|---|---|
| ***'Same-day/most recent' baseline covariates*** | | | | | |
| Sex | cov_cat_sex | Categorical | Male, Female | Primary care (patients) | Any other code than M and F is excluded by design (see qa_bin_is_female_or_male) |
| Age at baseline | cov_num_age | Continuous | | Primary care (patients) | Will be modelled using splines<br>Baseline: elig_date_t2dm |

| | | | | | |
|---|---|---|---|---|---|
| | derived: cov_cat_age | | "18-39", "40-59", "60-79", "80+" | | |
| Ethnicity | cov_cat_ethnicity | Categorical | White<br>Black<br>Asian<br>Mixed<br>Other<br>Unknown | Primary care (clinical_events) and SUS: ethnicity_snomed | Additionally included ethnicity_from_sus; codelist for primary care (snomed), based on recent publication: https://pmc.ncbi.nlm.nih.gov/articles/PMC11234682/<br>Based on entire EHR<br><br>Mixed, Other and Unknown was later collapsed |
| Index of Multiple Deprivation, at baseline | cov_cat_deprivation_5 | Categorical | 5 categories: 1 (most deprived) - 5 (least deprived) | Primary care (addresses) | Based on the Index of Multiple Deprivation score: https://docs.opensafely.org/legacy/study-def-variables/#cohortextractor.patients.address_as_of<br>Baseline: pandemicstart_date for Target Trial 1 (SeqTrial) and elig_date_t2dm for Target Trial 2 (modified Landmark)<br>https://data.cdrc.ac.uk/dataset/index-multiple-deprivation-imd<br>The IMD combines multiple aspects of deprivation into an overall score. These aspects include:<br>- Household overcrowding<br>- Homelessness<br>- Housing affordability<br>- Distance to key amenities like schools, supermarkets, and GP surgeries |
| Region, at baseline | strat_cat_region | Categorical | East<br>London<br>Midlands<br>North East and Yorkshire<br>North West<br>South East<br>South West | Primary care (practice_registrations) | Based on the Sustainable and Transformation Partnerships (STP) names and codes:<br>https://geoportal.statistics.gov.uk/documents/bec635f6c83e4582bcf76ce02c2be840/about<br>https://docs.opensafely.org/legacy/study-def-variables/#cohortextractor.patients.registered_practice_as_of<br>Baseline: elig_date_t2dm |
| Rural/urban classification, at baseline | cov_cat_rural_urban | Categorical | Urban conurbation<br>Urban city or town<br>Rural town or village | Primary care (addresses) | Baseline: elig_date_t2dm |
| Smoking status, on or before (most recent) baseline | cov_cat_smoking_status | Categorial | Smoker<br>Ever<br>Never<br>Unknown/Missing | Primary care (clinical_events): A combination of Smoking_clear & Ever_smoking | Baseline: elig_date_t2dm<br><br>Unknown/Missing and Ever later collapsed |
| Participant is a healthcare worker | cov_bin_healthcare_worker | Binary | T, F | NHS England COVID-19 data store (see: https://docs.opensafely.org/study-def-variables/#cohortextractor.patients.with_heal | Based on vaccination data, connected to the day of receiving a vaccination => at date in the future, but still a good proxy to use, since this status is long-term |

| | | | | | |
|---|---|---|---|---|---|
| | | | | thcare_worker_flag_on_covid_vaccine_record) | |
| Consultation rate, in year prior to baseline | cov_num_consrate | Numeric | Number of primary care contacts | Primary care | # QC: restrict to max 365 (average of one per day)<br>Baseline: elig_date_t2dm |
| BMI value, on or before (most recent) baseline (max. 2 years back) | cov_cat_bmi_groups;<br><br>cov_num_bmi | Categorical;<br><br>Numeric | 24 or lower or missing; 25-29; 30+<br><br>continuous | Primary care (clinical_events) | Biologically implausible BMI < 12 and > 70 replace with NA<br>Baseline: elig_date_t2dm |
| HbA1c value, on or before (most recent) baseline (max. 2 years back) | cov_cat_hba1c_mmol_mol<br><br>cov_num_hba1c_mmol_mol | Categorical<br><br>Numeric | <42; 42-58; 59-75; 75+; missing<br><br>Continuous (log-transformed) | Primary care (clinical_events):<br>hba1c_snomed | Implausible values set to missing:<br>< 0 or > 115<br>Categories based on<br>https://www.southtees.nhs.uk/resources/the-hba1c-test/<br>Baseline: elig_date_t2dm |
| Number of HbA1c measurements in previous 2 years | cov_num_counthba1c | Numeric | continuous | Primary care (clinical_events):<br>hba1c_measurement_snomed | |
| Total cholesterol/high-density lipoprotein [HDL] cholesterol ratio [TC/HDL], on or before (most recent) baseline (max. 2 years back) | cov_cat_tc_hdl_ratio<br><br>cov_num_tc_hdl_ratio | Categorical<br><br>Numeric | "below 3.5:1"; "3.5:1 to 5:1"; "above 5:1"; missing<br><br>continuous | Primary care (clinical_events)<br>cholesterol_snomed<br>hdl_cholesterol_snomed | TC/HDL values are derived from the recorded total and HDL cholesterol values.<br>Implausible values set to missing:<br>a) total Cholesterol values < 1.75 or > 20<br>b) HDL values < 0.4 or > 5<br>c) ratio values < 1 or > 50<br>Reference values/limits: https://doi.org/10.1093/ije/dyz099<br><br>https://www.urmc.rochester.edu/encyclopedia/content?ContentTypeID=167&ContentID=lipid_panel_hdl_ratio#:~:text=Most%20healthcare%20providers%20want%20the,1%20is%20considered%20very%20good.<br><br>Baseline: elig_date_t2dm |
| ***'Ever' baseline covariates (<u>any history</u> of)*** | | | | | |
| Obesity, on or before baseline | cov_bin_obesity | Binary | T, F | 1) Primary care (clinical_events):<br>- bmi_obesity_snomed_clinical<br>2) HES APC (apcs):<br>- bmi_obesity_icd10 | Baseline: elig_date_t2dm<br><br>Later in pipeline combined with cov_cat_bmi_groups mentioned above |

| | | | | | |
|---|---|---|---|---|---|
| Acute myocardial infarction, on or before baseline | cov_bin_ami | Binary | T, F | 1) Primary care (clinical_events):<br>- ami_snomed_clinical<br>2) HES APC (apcs):<br>- ami_prior_icd10<br>- ami_icd10 | Baseline: elig_date_t2dm |
| All stroke, on or before baseline | cov_bin_all_strok e | Binary | T, F | 1) Primary care (clinical_events):<br>- stroke_isch_snomed_clinical<br>- stroke_sah_hs_snomed_clinical<br>2) HES APC (apcs):<br>- stroke_isch_icd10<br>- stroke_sah_hs_icd10 | Baseline: elig_date_t2dm |
| Other arterial embolism, on or before baseline | cov_bin_other_art erial_embolism | Binary | T, F | 1) Primary care (clinical_events):<br>- other_arterial_embolism_snomed_clinical<br>2) HES APC (apcs):<br>- other_arterial_embolism_icd10 | Baseline: elig_date_t2dm |
| Venous thromboembolism events, on or before baseline | cov_bin_vte | Binary | T, F | 1) Primary care *(clinical_events):*<br>- portal_vein_thrombosis_snomed_clinical, dvt_dvt_snomed_clinical, dvt_icvt_snomed_clinical, dvt_pregnancy_snomed_clinical, other_dvt_snomed_clinical, pe_snomed_clinical<br><br>2) HES APC *(apcs):*<br>- portal_vein_thrombosis_icd10, dvt_dvt_icd10, dvt_icvt_icd10, dvt_pregnancy_icd10, other_dvt_icd10, icvt_pregnancy_icd10, pe_icd10 | Baseline: elig_date_t2dm |
| Heart failure, on or before baseline | cov_bin_hf | Binary | T, F | 1) Primary care (clinical_events):<br>hf_snomed_clinical<br>2) HES APC (apcs):<br>hf_icd10 | Baseline: elig_date_t2dm |
| Angina, on or before baseline | cov_bin_angina | Binary | T, F | 1) Primary care (clinical_events):<br>angina_snomed_clinical<br>2) HES APC (apcs):<br>angina_icd10 | Baseline: elig_date_t2dm |

| | | | | | |
|---|---|---|---|---|---|
| Dementia, on or before baseline | cov_bin_dementia | Binary | T, F | 1) Primary care (clinical_events): dementia_snomed_clinical dementia_vascular_snomed_clinical 2) HES APC (apcs): dementia_icd10 dementia_vascular_icd10 | Baseline: elig_date_t2dm |
| Cancer, on or before baseline | cov_bin_cancer | Binary | T, F | 1) Primary care (clinical_events): cancer_snomed_clinical 2) HES APC (apcs): cancer_icd10 | Baseline: elig_date_t2dm |
| Hypertension, on or before baseline | cov_bin_hypertension | Binary | T, F | 1) Primary care (clinical_events): hypertension_snomed_clinical 2) HES APC (apcs): hypertension_icd10 | Baseline: elig_date_t2dm |
| Depression, on or before baseline | cov_bin_depression | Binary | T, F | 1) Primary care (clinical_events): depression_snomed_clinical 2) HES APC (apcs): depression_icd10 | Baseline: elig_date_t2dm. |
| Chronic obstructive pulmonary disease , on or before baseline | cov_bin_copd | Binary | T, F | 1) Primary care (clinical_events): copd_snomed_clinical 2) HES APC (apcs): copd_icd10 | Baseline: elig_date_t2dm |
| Liver disease, on or before baseline | cov_bin_liver_disease | Binary | T, F | 1) Primary care (clinical_events): liver_disease_snomed_clinical 2) HES APC (apcs): liver_disease_icd10 | Baseline: elig_date_t2dm |
| Chronic kidney disease, on or before baseline | cov_bin_chronic_kidney_disease | Binary | T, F | 1) Primary care (clinical_events): ckd_snomed_clinical 2) HES APC (apcs): ckd_icd10 | Baseline: elig_date_t2dm |
| Polycystic Ovary Syndrome, on or before baseline | cov_bin_pcos | Binary | T, F | 1) Primary care (clinical_events): pcos_snomed_clinical 2) HES APC (apcs): pcos_icd10 | Baseline: elig_date_t2dm |
| Prediabetes, on or before baseline | cov_bin_prediabetes | Binary | T, F | Primary care (clinical_events): - prediabetes_snomed or - hba1c_new_codes, where((clinical_events.numeric_value>=42) & (clinical_events.numeric_value<=47.9)) | preDM HbA1c measure in period before index_date in preDM range (mmol/mol): 42-47.9. Baseline: elig_date_t2dm |

| | | | | | |
|---|---|---|---|---|---|
| Diabetes-related complications (Diab. Foot/Neuro/Nephro/Ret), on or before baseline | cov_bin_diabetescomp | Binary | T, F | 1) Primary care (clinical_events): diabetescomp_snomed_clinical<br>2) HES APC (apcs): diabetescomp_icd10 | Baseline: elig_date_t2dm |

### TABLES

**Appendix Table S1:** Exposure/treatment variables

| Variable | Name | Type | Definition | Data sources (tpp schema in brackets) | Notes |
|---|---|---|---|---|---|
| Metformin prescription | exp_bin_metfin | Binary | T, F | Primary care (medications):<br>- metformin_mono_dmd | First Metformin monotherapy prescription before studyend_date. Define/assign treatment strategy in R. |
| No antidiabetic prescription | exp_bin_treat_nothing | Binary | T, F | Primary care (medications), none of these:<br>- metformin_dmd<br>- sulfonylurea_dmd<br>- dpp4_dmd<br>- tzd_dmd<br>- sglt2_dmd<br>- glp1_dmd<br>- meglitinides_dmd<br>- agi_dmd<br>- insulin_dmd | First antidiabetic prescription before studyend_date. Define/assign treatment strategy in R. |
| On DM diet intervention only | cov_bin_diet_only | Binary | T, F | Primary care (clinical_events):<br>- diet_only_snomed | First code before studyend_date |

**Appendix Table S2:** Outcome variables

| Variable | Name | Type | Definition | Data sources (*tpp schema in brackets*) | Notes |
|---|---|---|---|---|---|
| First COVID-19 related hospitalisation or death, after baseline | out_date_severecovid | Date | First/any covid-19 (emergency) hospitalisation or covid-19 death | 1) HES APC (apcs): ["U071", "U072", "U109", "U099", "U074", "U075", "U10", "U09"]<br>2) Emergency care (emergency_care_attendances): ["1240751000000100", "1325171000000109", "1325181000000106"]<br>3) Death registry from ONS (ons_death): ["U071", "U072", "U109", "U099", "U074", "U075", "U10", "U09"] | Including clinically diagnosed COVID-19.<br><br>Search in all 25 emergency diagnosis fields.<br><br>Search in underlying_cause_of_death as well as in all other 15 cause_of_death fields |
| First COVID-19 event diagnosis, after baseline | out_date_covid | Date | First covid-19 event | 1) Primary care *(clinical_events):*<br>- covid_primary_care_code<br>- covid_primary_care_positive_test<br>- covid_primary_care_sequelae<br>2) SGSS *(sgss_covid_all_tests)*<br>3) HES APC *(apcs):*<br>["U071", "U072", "U109", "U099", "U074", "U075", "U10", "U09"]<br>4) Emergency care (emergency_care_attendances): covid_emergency -> ["1240751000000100", "1325171000000109", "1325181000000106"] | SGSS: COVID-19 tests results from SGSS (the Second Generation Surveillance System)<br><br>HES APC: including clinically diagnosed covid-19.<br><br>Adaptation based on:<br>https://github.com/opensafely/comparative-booster-spring2023/blob/main/analysis/codelists.py |
| First Long COVID diagnosis, after baseline | out_date_longvcovid_virfat | Binary; Date | First/any recorded Long COVID diagnosis | 1) Primary care (clinical_events):<br>long_covid_diagnostic_codes<br>long_covid_referral_codes<br>long_covid_assessment_codes<br>post_viral_fatigue_codes<br>2) HES APC (apcs):<br>["U109", "U099", "U074", "U075", "U10", "U09"] | Including post viral fatigue codes, see current long covid definition/repo here |
| De-registration date, after baseline | cens_date_dereg | Date | Deregistration date from TPP | Primary care (practice_registrations) | |
| *Sensitivity analyses, negative & positive outcome control:* In ehrQL: search for **first code** after baseline, then apply outcome window in R | | | | | |
| Hospitalisation due to fracture, after landmark/pandemic start | out_date_fracture | Date | First/any diagnosis, after pandemic start | 1) HES APC (apcs):<br>fracture_icd10 | |
| Diabetes-related death, after landmark/pandemic start | out_date_dm_death | Date | First/any diagnosis, after pandemic start | Death registry from ONS (ons_death):<br>diabetes_type2_icd10 | |

**Appendix Table S3:** Characteristics of people who died or loss to follow-up until landmark

| Characteristic | Alive and in care at landmark | Died or LTFU until landmark |
|---|---|---|
| | 52341 | 1377 |
| **Age, median (IQR), mean (SD)** | 62 (52, 71); 61 (13) | 64 (53, 75); 63 (15) |
| **Age groups** | | |
| 18-39 | 2,877 (5.5%) | 93 (6.8%) |
| 40-59 | 20,211 (38.6%) | 459 (33.3%) |
| 60-79 | 25,113 (48%) | 609 (44.2%) |
| 80 or older | 4,131 (7.9%) | 213 (15.5%) |
| **Sex** | | |
| Female | 23,901 (45.7%) | 573 (41.6%) |
| Male | 28,437 (54.3%) | 801 (58.2%) |
| **Ethnicity** | | |
| White | 41,919 (80.1%) | 1,131 (82.1%) |
| Asian | 6,861 (13.1%) | 141 (10.2%) |
| Black | 1,557 (3%) | 45 (3.3%) |
| Other | 2,007 (3.8%) | 63 (4.6%) |
| **Deprivation** | | |
| 1 (most deprived) | 12,807 (24.5%) | 357 (25.9%) |
| 2 | 11,631 (22.2%) | 297 (21.6%) |
| 3 | 11,187 (21.4%) | 291 (21.1%) |
| 4 | 9,159 (17.5%) | 231 (16.8%) |
| 5 (least deprived) | 7,545 (14.4%) | 189 (13.7%) |
| **Region** | | |
| East | 11,061 (21.1%) | 297 (21.6%) |
| London | 4,377 (8.4%) | 111 (8.1%) |
| Midlands | 10,725 (20.5%) | 273 (19.8%) |
| North East and Yorkshire | 10,923 (20.9%) | 279 (20.3%) |
| North West | 5,427 (10.4%) | 123 (8.9%) |
| South East | 3,477 (6.6%) | 99 (7.2%) |
| South West | 6,345 (12.1%) | 195 (14.2%) |
| **Rural/urban** | | |
| Urban conurbation | 16,629 (31.8%) | 405 (29.4%) |
| Urban city or town | 25,569 (48.9%) | 705 (51.2%) |
| Rural town or village | 10,137 (19.4%) | 261 (19%) |
| **Smoking status** | | |
| Smoker | 8,481 (16.2%) | 261 (19%) |
| Ever | 23,439 (44.8%) | 657 (47.7%) |
| Never | 20,415 (39%) | 453 (32.9%) |

| | | |
|---|---|---|
| **Healthcare worker** | 1,155 (2.2%) | 21 (1.5%) |
| **Consultation rate in previous year, median (IQR), mean (SD)** | 8 (5, 14); 11 (9) | 9 (5, 16); 12 (11) |
| **Body Mass Index (BMI) categories** | | |
| Underweight, healthy, or unknown | 17,157 (32.8%) | 549 (39.9%) |
| Overweight (25-29.9) | 10,161 (19.4%) | 285 (20.7%) |
| Obese (>30) | 25,023 (47.8%) | 543 (39.4%) |
| **HbA1c in mmol/mol** | 50 (48, 54); 52 (7) | 50 (48, 55); 52 (7) |
| **HbA1c categories in mmol/mol** | | |
| below 42 | 1,575 (3%) | 63 (4.6%) |
| 42-58 | 43,509 (83.1%) | 1,119 (81.3%) |
| 59-75 | 7,251 (13.9%) | 201 (14.6%) |
| **TC/Chol ratio** | 4.00 (3.25, 4.92); 4.19 (1.32) | 3.92 (3.07, 5.00); 4.14 (1.42) |
| **TC/HDL ratio categories** | | |
| below 3.5:1 | 13,953 (26.7%) | 399 (29%) |
| 3.5:1 to 5:1 | 19,401 (37.1%) | 441 (32%) |
| above 5:1 | 9,087 (17.4%) | 249 (18.1%) |
| Unknown | 9,897 (18.9%) | 279 (20.3%) |
| **History of acute myocardial infarct** | 4,245 (8.1%) | 141 (10.2%) |
| **History of stroke** | 1,983 (3.8%) | 75 (5.4%) |
| **History of other arterial embolism** | 171 (0.3%) | 9 (0.7%) |
| **History of venous thromboembolism** | 1,743 (3.3%) | 57 (4.1%) |
| **History of heart failure** | 2,859 (5.5%) | 147 (10.7%) |
| **History of angina pectoris** | 6,069 (11.6%) | 207 (15%) |
| **History of dementia** | 645 (1.2%) | 57 (4.1%) |
| **History of cancer** | 14,985 (28.6%) | 411 (29.8%) |
| **History of arterial hypertension** | 27,471 (52.5%) | 747 (54.2%) |
| **History of depression** | 18,525 (35.4%) | 531 (38.6%) |
| **History of COPD** | 4,695 (9%) | 213 (15.5%) |
| **History of liver disease** | 393 (0.8%) | 9 (0.7%) |
| **History of CKD** | 5,661 (10.8%) | 261 (19%) |
| **History of PCOS** | 51 (0.1%) | 3 (0.2%) |
| **History of prediabetes** | 36,303 (69.4%) | 939 (68.2%) |
| **Diabetes complication** | 447 (0.9%) | 9 (0.7%) |
| **Count HbA1c measurements, previous 2y, median (IQR), mean (SD)** | 3.00 (2.00, 4.00); 3.06 (1.87) | 3.00 (2.00, 4.00); 3.07 (2.02) |
| **Count lifestyle advices, previous 2y, median (IQR), mean (SD)** | 0.00 (0.00, 1.00); 0.61 (1.15) | 0.00 (0.00, 1.00); 0.61 (1.57) |

**Appendix Table S4:** Characteristics of people who died or loss to follow-up until pandemic start

| Characteristic | Alive and in care at pandemic start | Died or LTFU between landmark and pandemic start |
|---|---|---|
| | 50991 | 1347 |
| **Age, median (IQR), mean (SD)** | 62 (52, 71); 61 (13) | 64 (51, 75); 63 (15) |
| **Age groups** | | |
| 18-39 | 2,775 (5.4%) | 105 (7.8%) |
| 40-59 | 19,791 (38.8%) | 423 (31.4%) |
| 60-79 | 24,483 (48%) | 627 (46.5%) |
| 80 or older | 3,939 (7.7%) | 189 (14%) |
| **Sex** | | |
| Female | 23,289 (45.7%) | 609 (45.2%) |
| Male | 27,705 (54.3%) | 735 (54.6%) |
| **Ethnicity** | | |
| White | 40,845 (80.1%) | 1,071 (79.5%) |
| Asian | 6,681 (13.1%) | 177 (13.1%) |
| Black | 1,515 (3%) | 39 (2.9%) |
| Other | 1,941 (3.8%) | 63 (4.7%) |
| **Deprivation** | | |
| 1 (most deprived) | 12,459 (24.4%) | 345 (25.6%) |
| 2 | 11,319 (22.2%) | 309 (22.9%) |
| 3 | 10,917 (21.4%) | 273 (20.3%) |
| 4 | 8,913 (17.5%) | 243 (18%) |
| 5 (least deprived) | 7,371 (14.5%) | 171 (12.7%) |
| **Region** | | |
| East | 10,737 (21.1%) | 327 (24.3%) |
| London | 4,257 (8.3%) | 117 (8.7%) |
| Midlands | 10,461 (20.5%) | 261 (19.4%) |
| North East and Yorkshire | 10,671 (20.9%) | 249 (18.5%) |
| North West | 5,301 (10.4%) | 129 (9.6%) |
| South East | 3,405 (6.7%) | 75 (5.6%) |
| South West | 6,153 (12.1%) | 189 (14%) |
| **Rural/urban** | | |
| Urban conurbation | 16,209 (31.8%) | 417 (31%) |
| Urban city or town | 24,897 (48.8%) | 675 (50.1%) |
| Rural town or village | 9,885 (19.4%) | 255 (18.9%) |
| **Smoking status** | | |
| Smoker | 8,229 (16.1%) | 249 (18.5%) |
| Ever | 22,815 (44.7%) | 627 (46.5%) |
| Never | 19,947 (39.1%) | 471 (35%) |
| **Healthcare worker** | 1,137 (2.2%) | 21 (1.6%) |
| **Consultation rate in previous year, median (IQR), mean (SD)** | 8 (5, 13); 11 (9) | 10 (6, 16); 13 (11) |
| **Body Mass Index (BMI) categories** | | |
| Underweight, healthy, or unknown | 16,647 (32.6%) | 507 (37.6%) |

| | | |
|---|---|---|
| Overweight (25-29.9) | 9,885 (19.4%) | 279 (20.7%) |
| Obese (>30) | 24,459 (48%) | 561 (41.6%) |
| **HbA1c in mmol/mol** | 50 (48, 54); 52 (7) | 50 (48, 54); 52 (7) |
| **HbA1c categories in mmol/mol** | | |
| below 42 | 1,515 (3%) | 63 (4.7%) |
| 42-58 | 42,429 (83.2%) | 1,077 (80%) |
| 59-75 | 7,047 (13.8%) | 207 (15.4%) |
| **TC/Chol ratio** | 4.00 (3.25, 4.92); 4.19 (1.32) | 3.97 (3.18, 4.88); 4.13 (1.33) |
| **TC/HDL ratio categories** | | |
| below 3.5:1 | 13,587 (26.6%) | 363 (26.9%) |
| 3.5:1 to 5:1 | 18,927 (37.1%) | 477 (35.4%) |
| above 5:1 | 8,871 (17.4%) | 213 (15.8%) |
| Unknown | 9,603 (18.8%) | 291 (21.6%) |
| **History of acute myocardial infarct** | 4,119 (8.1%) | 123 (9.1%) |
| **History of stroke** | 1,905 (3.7%) | 75 (5.6%) |
| **History of other arterial embolism** | 165 (0.3%) | 9 (0.7%) |
| **History of venous thromboembolism** | 1,677 (3.3%) | 63 (4.7%) |
| **History of heart failure** | 2,745 (5.4%) | 117 (8.7%) |
| **History of angina pectoris** | 5,895 (11.6%) | 171 (12.7%) |
| **History of dementia** | 591 (1.2%) | 51 (3.8%) |
| **History of cancer** | 14,589 (28.6%) | 393 (29.2%) |
| **History of arterial hypertension** | 26,751 (52.5%) | 717 (53.2%) |
| **History of depression** | 18,003 (35.3%) | 525 (39%) |
| **History of COPD** | 4,491 (8.8%) | 201 (14.9%) |
| **History of liver disease** | 381 (0.7%) | 9 (0.7%) |
| **History of CKD** | 5,481 (10.7%) | 177 (13.1%) |
| **History of PCOS** | 45 (0.1%) | 3 (0.2%) |
| **History of prediabetes** | 35,403 (69.4%) | 903 (67%) |
| **Diabetes complication** | 441 (0.9%) | 9 (0.7%) |
| **Count HbA1c measurements, previous 2y, median (IQR), mean (SD)** | 3.00 (2.00, 4.00); 3.06 (1.86) | 3.00 (2.00, 4.00); 3.12 (2.08) |
| **Count lifestyle advices, previous 2y, median (IQR), mean (SD)** | 0.00 (0.00, 1.00); 0.61 (1.15) | 0.00 (0.00, 1.00); 0.58 (1.19) |

**Appendix Table S5.** 2-year risk estimates for the primary outcome

| | **Cumulative risk in metformin group (95% CI)** | **Cumulative risk in no antidiabetic group (95% CI)** | **Adjusted risk ratio (95% CI)** | **Adjusted risk difference (95% CI)** | **Adjusted hazard ratio (95% CI)** |
|---|---|---|---|---|---|
| **Intention to treat** | 2.23% (2.02-2.45)* | 1.87% (1.72-2.03)* | 1.19 (1.05-1.35)* | 0.36% (0.09-0.62)* | 1.17 (1.02-1.35)° |
| **Per protocol** | 2.14% (1.92-2.36)# | 1.93% (1.72-2.14)# | 1.11 (0.96-1.29)# | 0.21% (-0.08-0.51)# | 1.11 (0.96-1.29)§ |
| **Per protocol,** untruncated weights | 2.16% (1.93-2.40)# | 1.92% (1.72-2.12)# | 1.13 (0.97-1.31)# | 0.24% (-0.06-0.54)# | 1.13 (0.97-1.32)§ |

Footnote:
*from weighted Kaplan Meier
°from Cox model
#from pooled logistic regression with treatment-time interaction
§from pooled logistic regression without treatment-time interaction to model proportional hazards

#### FIGURES

**Appendix Figure S1:** Directed acyclic graph

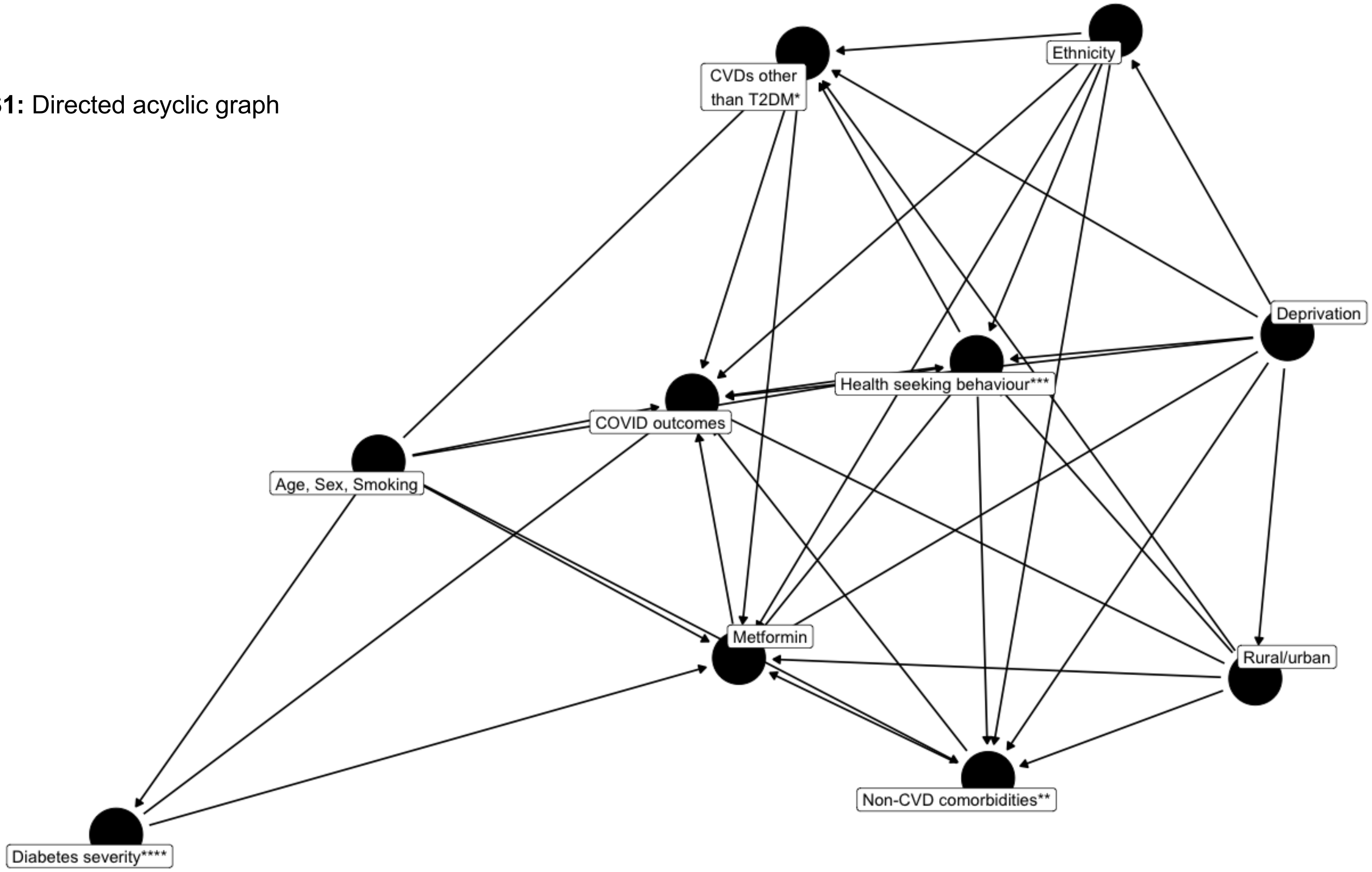


This DAG focuses on the primary outcome (severe COVID). We conduct region-stratified analyses, thus, region is not part of the DAG. It would have the same representation as “Rural/urban”.
Calendar time was an additional adjustment variable to control for older (mid 2018) vs newer (mid 2019) T2DM diagnoses
CVD = Cardiovascular disease
* CVDs other than T2DM: Total Chol/HDL ratio, preDM, PCOS, stroke, angina, acute myocardial infarction, other arterial embolism, venous thromboembolism events, heart failure, hypertension
** non-CVD comorbidities: dementia, cancer, chronic obstructive pulmonary disease, liver disease,
chronic kidney disease, depression
*** Health seeking behaviour; proxies used: Is a healthcare worker; number of primary care consultation rate in previous year, number of HbA1c measurements in prior 2 years
**** HbA1c levels and T2DM complications

**Appendix Figure S2.** Density plots (smoothed histogram), based on propensity score of receiving metformin

A. Total study population

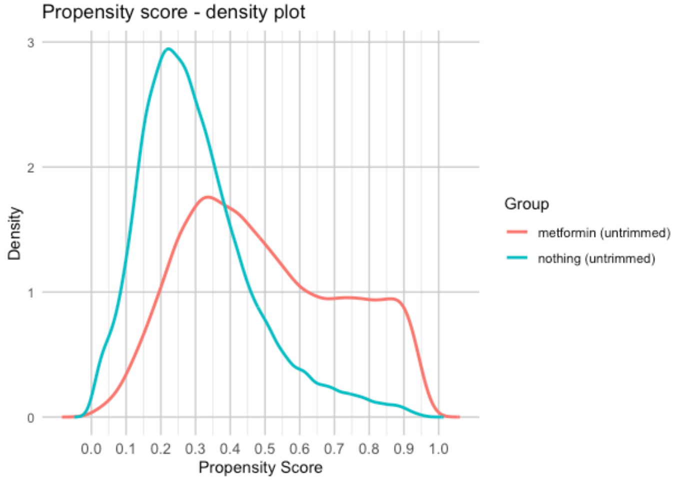


B. By HbA1c subgroup

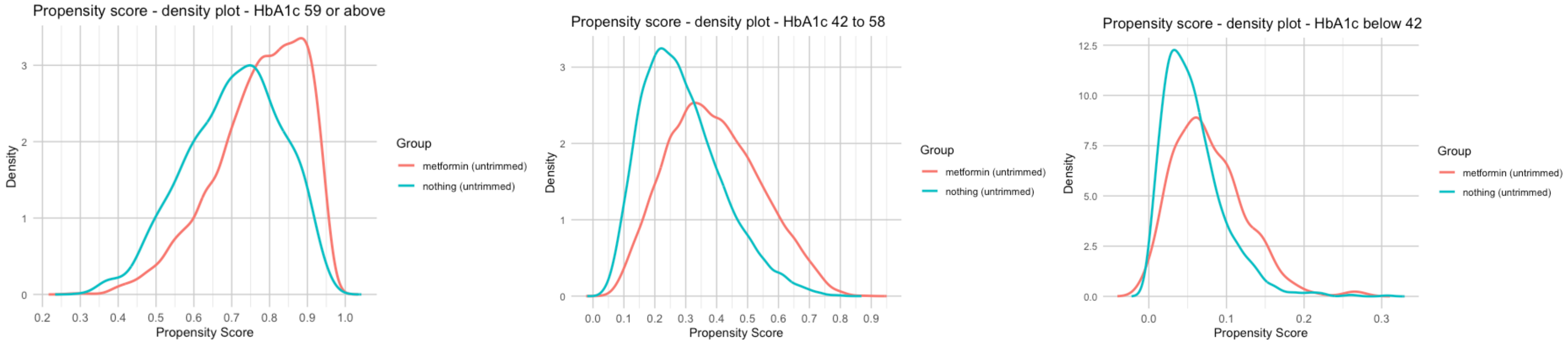

**Appendix Figure S3.** All outcomes, minimal adjusted model, primary analysis (Cox model, intention-to-treat)

| Outcome | Events Metformin n (%) | Events No Antidiabetic n (%) | aHR (95% CI) |
|---|---|---|---|
| Primary: COVID−19−related hospitalization or death | 375 (1.92%) | 597 (1.82%) | 1.18 (1.03 – 1.34) |
| Secondary: Any COVID−19 diagnosis | 1863 (9.52%) | 2565 (7.84%) | 1.08 (1.02 – 1.15) |
| Secondary: Long COVID | 33 (0.17%) | 33 (0.10%) | 1.41 (0.87 – 2.28) |
| Negative control outcome: Bone fracture | 159 (0.81%) | 351 (1.07%) | 1.05 (0.87 – 1.27) |
| Positive control outcome: Diabetes−related death | 123 (0.63%) | 195 (0.60%) | 1.58 (1.26 – 1.99) |
| Sensitivity: Propensity score overlap population | 327 (1.96%) | 549 (1.82%) | 1.16 (1.01 – 1.33) |

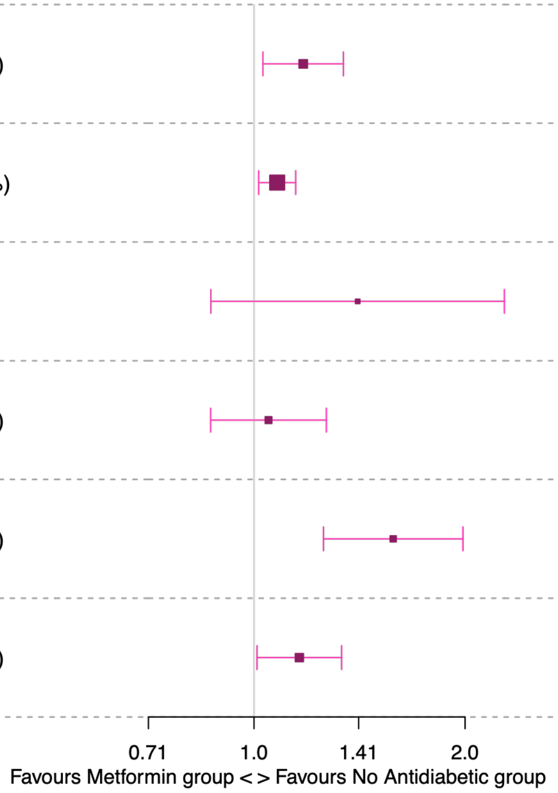


*Footnote: hazard ratio, adjusted for age and sex; abbreviations: aHR (adjusted Hazard Ratio), CI (confidence intervals)*

**Appendix Figure S4.** Primary outcome, cumulative incidence curves, weighted Kaplan Meier (intention-to-treat)

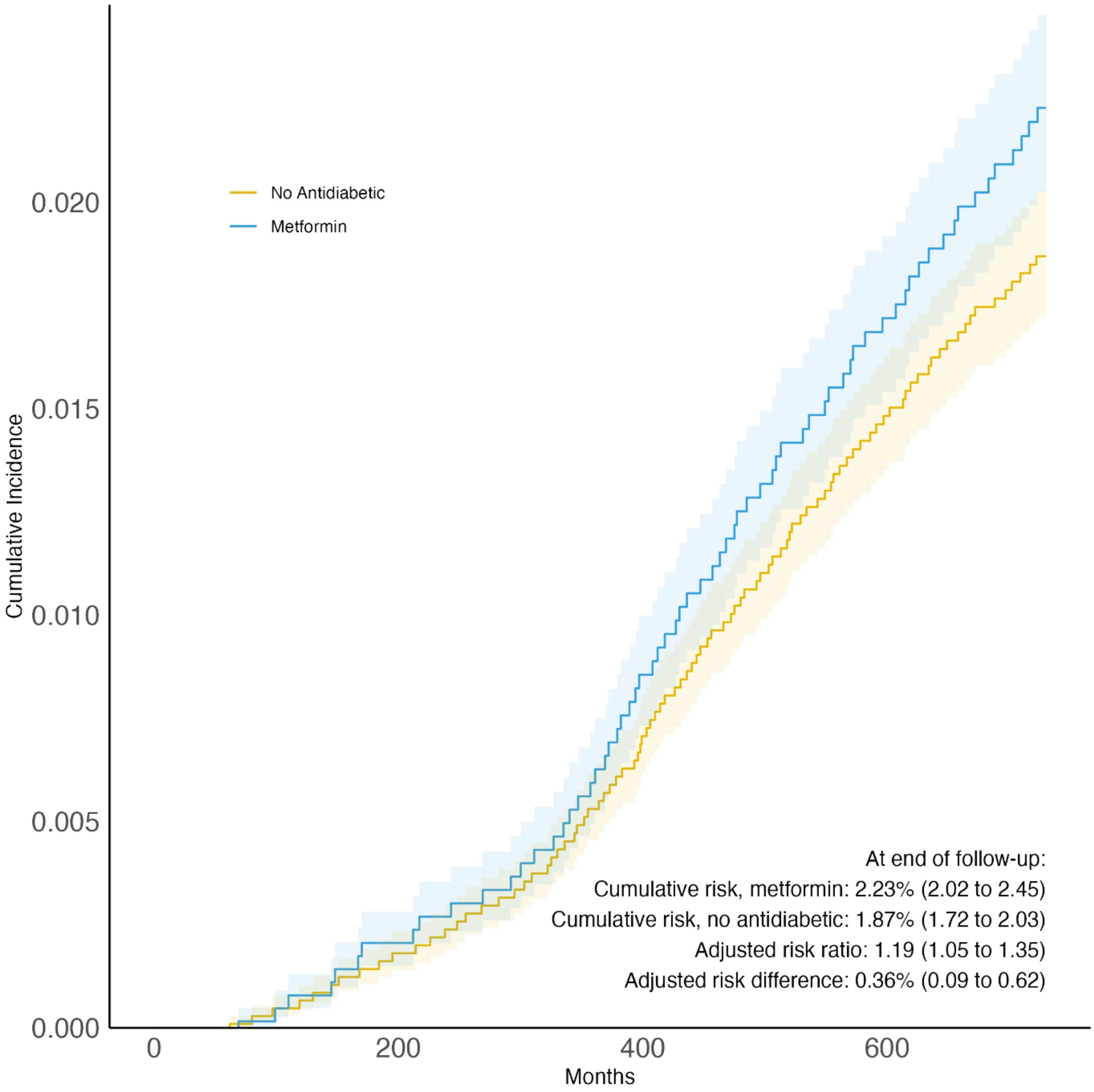

**Appendix Figure S5.** Primary outcome, cumulative incidence curves, pooled logistic regression (per-protocol)

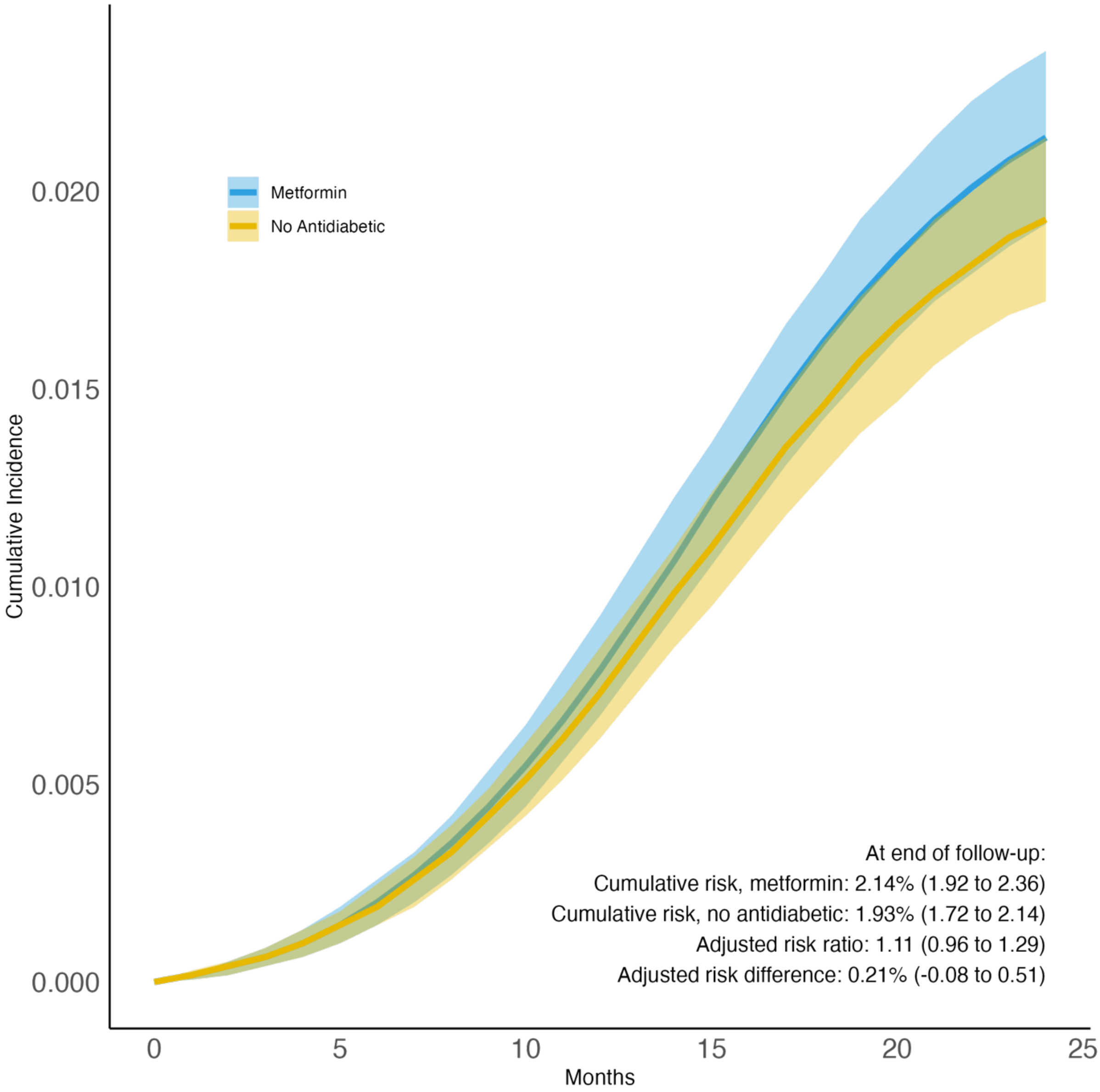

**Appendix Figure S6.** Forest plot, presenting subgroup analyses and tests for heterogeneity (Cochran's Q) on primary outcome

| Characteristic | Group | Events Metformin n (%) | Events No Antidiabetic n (%) | aHR (95% CI) | P–value for heterogeneity |
|---|---|---|---|---|---|
| **Age** | | | | | **0.106** |
| | 60 years or above | 213 (2.42%) | 411 (2.01%) | 1.29 (1.08 – 1.54) | |
| | below 60 years | 159 (1.48%) | 189 (1.54%) | 1.02 (0.81 – 1.28) | |
| **Sex** | | | | | **0.693** |
| | Female | 159 (1.78%) | 255 (1.70%) | 1.14 (0.91 – 1.41) | |
| | Male | 213 (2.00%) | 345 (1.94%) | 1.20 (1.00 – 1.45) | |
| **Ethnicity** | | | | | **0.283** |
| | Non–White | 87 (2.01%) | 135 (2.22%) | 1.04 (0.78 – 1.38) | |
| | White | 285 (1.87%) | 459 (1.72%) | 1.24 (1.05 – 1.46) | |
| **Deprivation** | | | | | **0.774** |
| | Most deprived | 135 (2.41%) | 177 (2.46%) | 1.14 (0.90 – 1.46) | |
| | Other | 237 (1.70%) | 417 (1.63%) | 1.19 (1.01 – 1.42) | |
| **BMI** | | | | | **<0.001** |
| | Obese | 195 (1.93%) | 327 (2.20%) | 0.92 (0.75 – 1.11) | |
| | Overweight | 51 (1.62%) | 105 (1.50%) | 1.37 (0.96 – 1.96) | |
| | Normal/low weight or missing | 129 (2.05%) | 159 (1.47%) | 1.78 (1.38 – 2.29) | |
| **HbA1c** | | | | | **0.283** |
| | below 59 mmol/mol | 285 (2.04%) | 579 (1.86%) | 1.15 (0.99 – 1.33) | |
| | 59 mmol/mol or above | 87 (1.56%) | 15 (0.91%) | 1.55 (0.91 – 2.65) | |

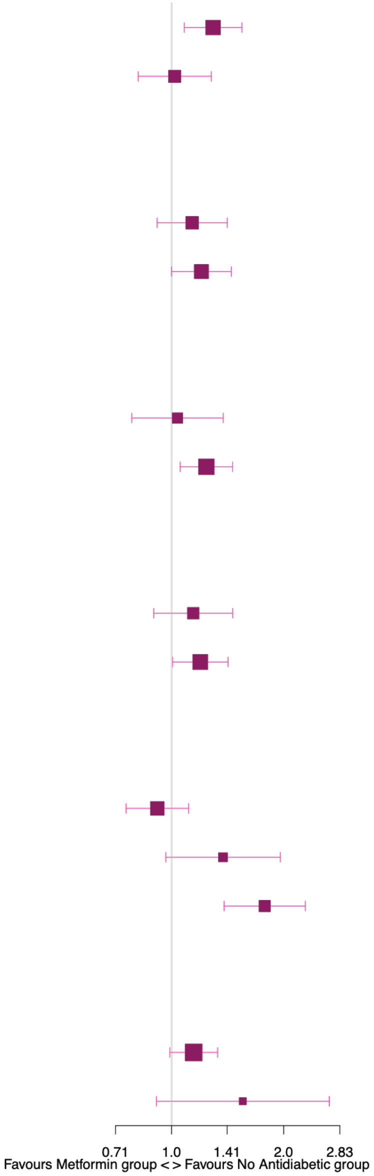
